# RATING: Medical-knowledge-guided rheumatoid arthritis assessment from multimodal ultrasound images via deep learning

**DOI:** 10.1101/2022.04.08.22273640

**Authors:** Zhanping Zhou, Chenyang Zhao, Hui Qiao, Ming Wang, Yuchen Guo, Qian Wang, Rui Zhang, Huaiyu Wu, Fajin Dong, Zhenhong Qi, Jianchu Li, Xinping Tian, Xiaofeng Zeng, Yuxin Jiang, Feng Xu, Qionghai Dai, Meng Yang

## Abstract

Ultrasound (US) examination has been commonly utilized in clinical practice for assessing the rheumatoid arthritis (RA) activity, which is hampered by low intra-observer and inter-observer agreement as well as considerable time and expense to train experienced radiologists. Here, we present the Rheumatoid ArthriTIs kNowledge Guided (RAT ING) model that scores RA activity and generates interpretable features to assist radiologists’ decision-making. The RATING model achieved an accuracy of 86.1% (95% confidence interval (CI)=82.5%–90.1%) in the clinical trial setting, and achieved an accuracy of 85.0% (95% CI=80.5%–89.1%) on the US images collected from an external medical center. An AI-assisted reader study demonstrated that the RATING model improved the average accuracy of ten radiologists from 41.4% to 64.0%. Automated AI models for the assessment of RA may facilitate US RA examination and provide support for clinical decision-making.

## Introduction

Rheumatoid arthritis (RA), a systemic and chronic inflammation that mainly affects small joints, has detrimental outcomes on both individuals and society, including increased disability and mortality [1]. According to a treatment-to-target strategy, quantitative assessment of disease activity has been deemed as the key for evaluating the alleviation of RA disease burden [2]. Due to its radiation-free, non-invasive and cost-effective characteristics, ultrasound (US) examination has been commonly utilized in clinical practice for assessing the RA activity [3], which usually comprises two principal modes, grey-scale US (GSUS) and Doppler US (either color or power Doppler). GSUS images are examined to investigate the morphological changes of synovial hypertrophy (SH), while Doppler US images are examined to detect synovial hypervascularity [4]. However, there was a long time with no agreement among radiologists on how to evaluate the GSUS and Doppler US measurements until the introduction of European League Against Rheumatisms–Outcomes Measures in Rheumatology Synovitis Scoring (EOSS) system [5]. EOSS system was proposed to standardize the assessment procedure for US examination, which emphasizes the importance of analyzing both GSUS and Doppler US images. It developed the 0-3 scale scoring methods for SH and vascularity evaluations, and recommended the combined score of SH and vascularity based on expert consensus to further enhance the reliability.

Despite of the establishment of EOSS system, the quantitative assessment of disease activity is still hampered by low intra-observer and inter-observer agreement during US examination [6]. Additionally, it requires considerable time and expense to train experienced radiologists in US diagnostics, and integrating both two US modes to employ the EOSS system further aggravates the problem [1]. As an emerging alternative solution, a type of artificial intelligence (AI) methods called deep learning [7] has been currently utilized in US imaging analysis [8] and has shown great potential in clinical diagnosis of breast cancer [9] [10], thyroid cancer [11], liver malignancy [12], biliary atresia [13], and assessment of cardiac function [14] [15]. For RA assessment, a few AI models have also been proposed to score the disease activity [16] [17], which use a cascade of convolutional neural networks to directly predict the combined score with only Doppler US images.

Although the feasibility of AI methods has been demonstrated, the clinical applicability of AI-assisted RA assessment has yet to materialize owing to two major limitations. Firstly, previous studies used only one US imaging mode (that is, either GSUS or Doppler US), deviating from the EOSS system and thus resulted in potential issues relevant to clinical acceptance and diagnostic accuracy. In the clinical procedure where the EOSS system is adopted, both the measurements of GSUS and Doppler US are indispensable and imply unique RA characteristics. Secondly, previous studies have not conducted the clinical comparison trials and examined to what extent their systems might actually help and improve clinical diagnostics. While the majority of studies compare the performance of AI models with humans, it is more likely that humans could collaborate with AI models in real-life medical practice [18].

To overcome the hurdle of RA assessment in clinical practice, we propose Rheumatoid ArthriTIs kNowledge Guided (RATING) model for scoring the RA activity. Inspired by the clinical US assessment procedure during which radiologists combine both GSUS and Doppler US information to evaluate SH, we design a GS-Doppler feature fusion network that comprehensively analyze the measurements of the two modes. Besides, to further follow the real assessment process in which multiple radiologists discuss and jointly consider SH and vascularity scores to reach an agreement on RA scoring, our method presents a MULTI-Task mUlti-moDel Ensemble framework named MULTITUDE to make the full use of multiple SH and vascularity score predictions by fusing them to obtain the final combined score. We collected 752 paired GSUS and Doppler US images from Peking Union Medical College Hospital (PUMCH) as our development dataset to train the RATING model. We demonstrate that the RATING model has the potential to offer reliable diagnosis, implements well in clinical trial settings, and generalizes well to different US operators and Doppler US modes. Furthermore, we build an assistance software that provides explainable features and AI predictions to assist radiologists’ decision-making. We demonstrate that the RATING model greatly improves radiologists’ scoring accuracy.

## Results

### Build of the RATING model on the development dataset

The over-all pipeline of building the RATING model is illustrated in Fig. 1. To build the model, we retrospectively collected a development dataset, which consists of 752 paired GSUS and color Doppler US (CDUS) images from 104 patients. For each pair of US images, three experienced radiologists from PUMCH reviewed the images, annotated the region of interest (ROI) for each US image, and decided the SH score, vascularity score and combined score. The workflow followed the EOSS guidelines (Supplementary Table 1) and is shown in Supplementary Fig. 1. Detailed patient demographics and EOSS scoring characteristics are summarized in Supplementary Table 2.

**Fig. 1.**
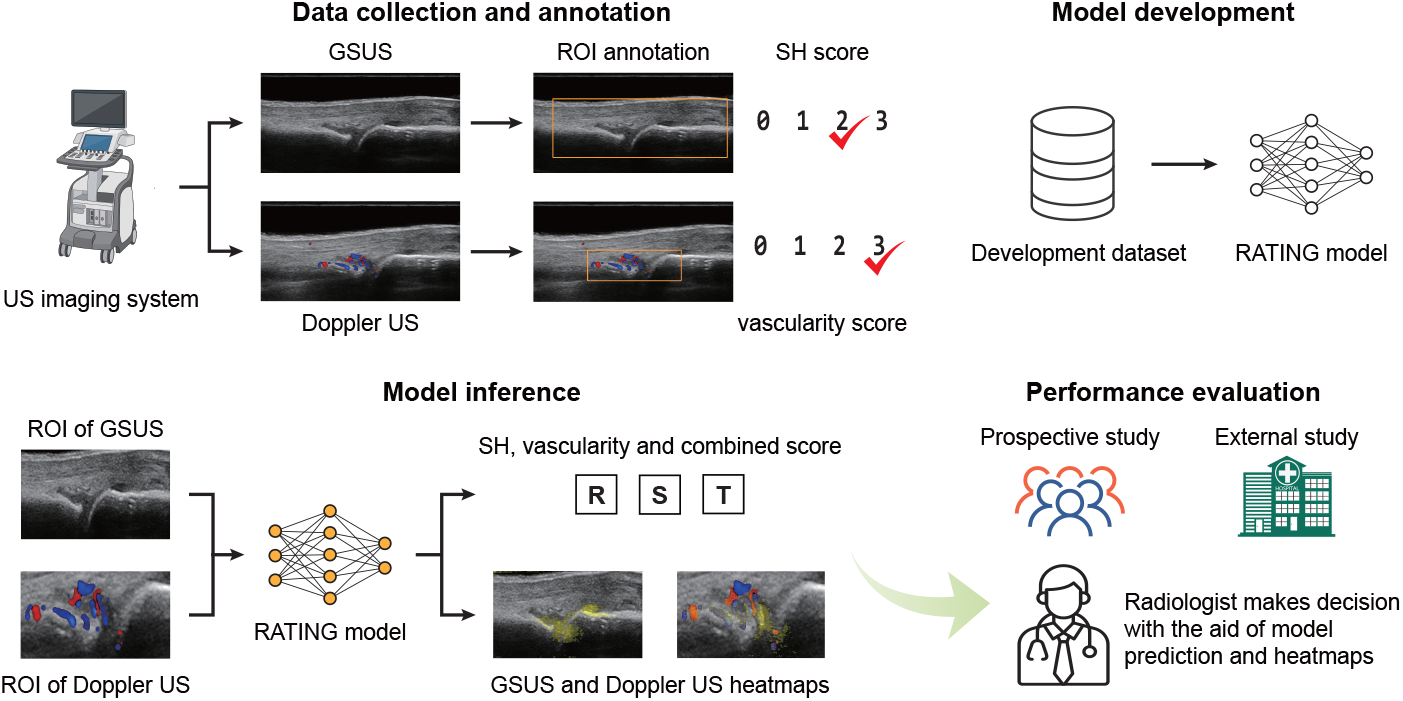
Build of the RATING model for RA scoring. Paired GSUS and Doppler US images were collected for the development dataset, the prospective test dataset and the external test dataset, then ROIs were annotated and scored according to the EOSS system. During model development, five models were trained separately on different datasets. For each pair of GSUS and Doppler US image, the RATING model predicts the SH score, the vascularity score and the combined score. Together with score predictions, heatmaps are generated for GSUS and Doppler US images and assist radiologists.

We trained the RATING model on the development dataset. The RATING model consists of five models which share the same architecture. Each of the five models separately predicts a SH score and a vascularity score. Instead of directly predicting the scores by multi-classification models, each model predicts three binary classification tasks, i.e., whether the SH score or the vascularity score is greater than 0, 1 and 2. During inference stage, for each sample, the five models separately predict the SH score and the vascularity score, and the combined score is predicted by comprehensively considering the above predictions by the MULTITUDE framework.

On three SH score binary classification tasks, the ROC curves on the development dataset are shown in Supplementary Fig. 3 and AUCs were 0.896 (95% confidence interval (CI)=0.883–0.909), 0.945 (95% CI=0.935– 0.956) and 0.948 (95% CI=0.932–0.964), respectively. Positive predictive value (PPV), negative predictive value (NPV), sensitivity and specificity are shown in Supplementary Table 3. On three vascularity score binary classification tasks, the ROC curves are shown in Supplementary Fig. 4 and AUCs were 0.980 (95% CI=0.976–0.984), 0.992 (95% CI=0.988–0.996) and 0.991 (95% CI=0.986–0.995), respectively. Detailed metrics are shown in Supplementary Table 4.

### Performance of the RATING model on the prospective test dataset

To evaluate the performance of the RATING model in clinical trial settings, 28 patients with RA were prospectively recruited from April 20th 2021 to October 2021 and received US examination at PUMCH. After radiologists reviewed and scored the US images, 274 paired GSUS and CDUS images were included in the prospective test dataset.

We evaluated the performance of the RATING model in two ways. Firstly, we calculated the area under the curve (AUC) of the receiver operating characteristic (ROC) curve for binary classification tasks, including three SH score binary classification tasks and three vascularity score binary classification tasks. Secondly, we calculated the four-class accuracy and linearly weighted *κ* [19] for SH score, vascularity score and combined score classification.

On three SH score binary classification tasks, the ROC curves on the prospective test dataset are shown in Supplementary Fig. 5 and AUCs were 0.930 (95% CI=0.919–0.941), 0.933 (95% CI=0.930–0.936) and 0.979 (95% CI=0.973–0.985), respectively. PPV, NPV, sensitivity and specificity are shown in Supplementary Table 5. On three vascularity score binary classification tasks, the ROC curves are shown in Supplementary Fig. 6 and AUCs were 0.986 (95% CI=0.985–0.987), 0.990 (95% CI=0.986–0.995) and 0.995 (95% CI=0.991–0.998), respectively. PPV, NPV, sensitivity and specificity are shown in Supplementary Table 6.

The RATING model achieved an accuracy score of 86.1% (95% CI=82.5%– 90.1%) for combined score prediction and a linearly weighted *κ* score of 0.853 (95% CI=0.806–0.900). The confusion matrix is shown in Supplementary Table 10, and Fig. 2a shows the normalized confusion matrix. For joints of combined score 0 and 4, the accuracy scores are higher than 90%. It seems to be most difficult for the RATING model to predict joints of combined score 1, which were frequently predicted as 0 and 2. As shown in Supplementary Table 7, the accuracy score of the SH score and the vascularity score were 79.6% (95% CI=74.8%–84.3%) and 94.5% (95% CI=91.6%–97.1%), and the linearly weighted *κ* score were 0.757 (95% CI=0.699–0.885) and 0.919 (95% CI=0.876– 0.966). Confusion matrices of the SH score classification and the vascularity score classification are shown in Supplementary Table 8 and 9, respectively.

**Fig. 2.**
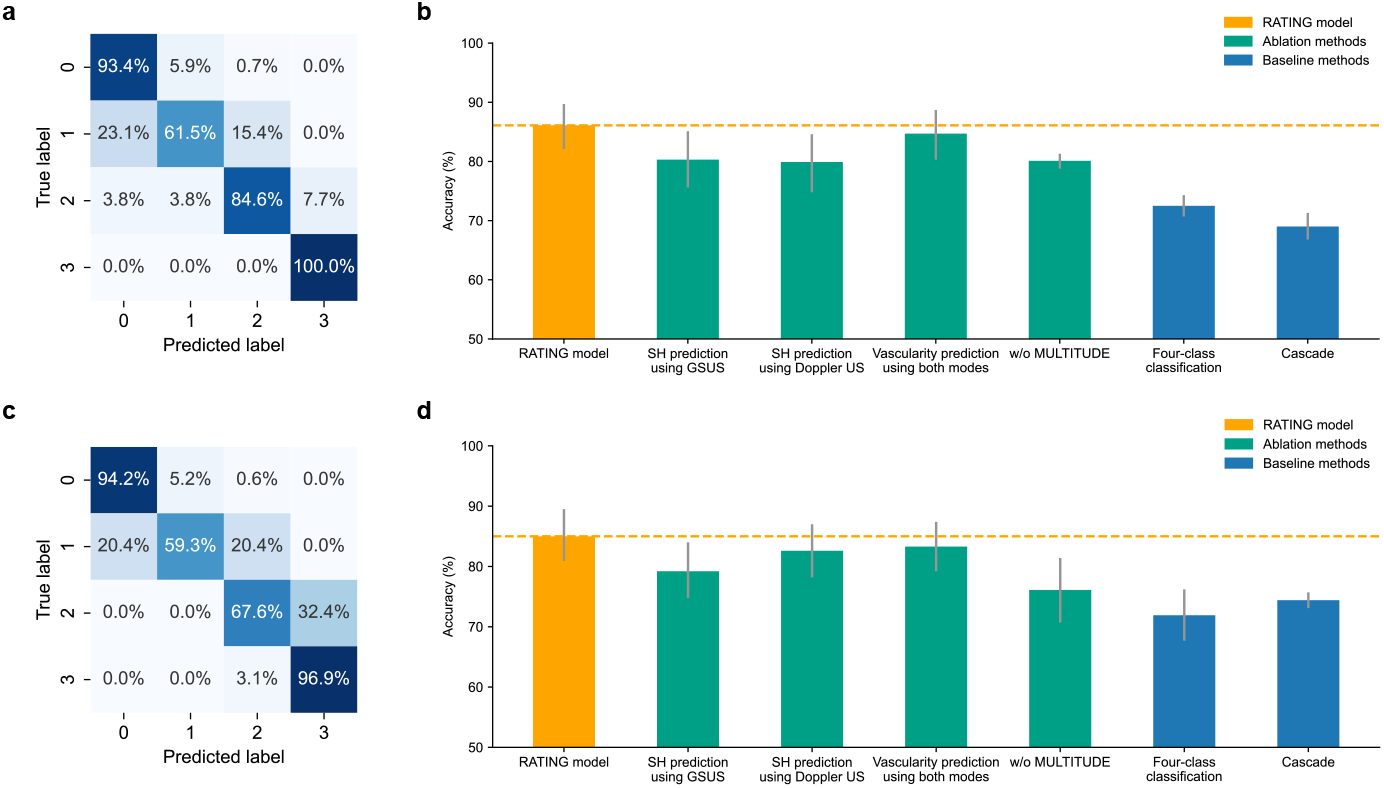
The performance of the RATING model in the classification of the combined score. Normalized confusion matrices (**a**,**c**) and accuracy comparison (**b**,**d**) for the combined score classification. **b**, The RATING model achieved accuracy=86.1%(95% CI=82.5%–90.1%) on the prospective test dataset, which was higher than ablation methods and baseline methods. **d**, The RATING model achieved accuracy=85.0%(95% CI=80.5%– 89.1%) on the external test dataset, which was higher than ablation methods and baseline methods. Error bars indicate 95% confidence intervals.

### Performance of the RATING model on the external test dataset

To further demonstrate the generalizability of the RATING model to different US operators and Doppler US modes, an external test dataset was collected from Shenzhen People’s Hospital (SZPH) from March 2021 to December 2021. Different from the development dataset and the prospective test dataset, the external test dataset consists of power Doppler US (PDUS) images instead of CDUS images. After radiologists reviewed and scored the US images, 315 paired GSUS and PDUS images from 42 patients were included in the dataset.

On three SH score binary classification tasks, the ROC curves on the external test dataset are shown in Supplementary Fig. 7 and AUCs were 0.940 (95% CI=0.920–0.960), 0.985 (95% CI=0.983–0.988) and 0.979 (95% CI=0.974–0.984), respectively. PPV, NPV, sensitivity and specificity are shown in Supplementary Table 11. On three vascularity score binary classification tasks, the ROC curves are shown in Supplementary Fig. 8 and AUCs were 0.998 (95% CI=0.995–1.000), 0.996 (95% CI=0.994–0.998) and 0.988 (95% CI=0.974–1.000), respectively. PPV, NPV, sensitivity and specificity are shown in Supplementary Table 12.

The RATING model achieved an accuracy score of 85.0% (95% CI=80.5%– 89.1%) for combined score prediction and a linearly weighted *κ* score of 0.857 (95% CI=0.817–0.897). The confusion matrix is shown in Supplementary Table 16, and Fig. 2c shows the normalized confusion matrix. For joints of combined score 0 and 4, the accuracy scores are higher than 90%. Similar to the result on the prospective test dataset, it seems to be more difficult for the RATING model to predict joints of combined score 1 and 2. As shown in Supplementary Table 13, the accuracy score of the SH score and the vascularity score were 82.9% (95% CI=78.5%–87.0%) and 96.2% (95% CI=93.9%–98.3%), and the linearly weighted *κ* score were 0.832 (95% CI=0.789–0.919) and 0.957 (95% CI=0.932–0.953). Confusion matrices of the SH score classification and the vascularity score classification are shown in Supplementary Table 14 and 15, respectively. The experiment results demonstrate that the RATING model generalizes well to different US operators and Doppler US modes.

### Comparative studies of ablation methods and baseline methods

To assess the effectiveness of the GS-Doppler feature fusion network and the MULTITUDE framework, we evaluated the performance of ablation methods on the prospective test dataset and the external test dataset. For the GS-Doppler feature fusion network, we evaluated three ablation methods: (1) SH score predicted using only GSUS images, (2) SH score predicted using only Doppler US images, (3) vascularity score predicted using both GSUS and Doppler US images. For MULTITUDE, we evaluated the performance of the five single models. In addition, we evaluated two baseline methods on the prospective test dataset and the external test dataset. First, we replaced binary classification networks with four-class classification networks that directly predicted the SH score and the vascularity score. Second, we evaluated the cascade method that was proposed in a previous study [17].

On both the prospective test dataset (Fig. 2b) and the external test dataset (Fig. 2d), our methods achieved significant higher accuracy score and linearly weighted *κ* score with *P <* 0.001. The experiment results demonstrate the effectiveness of the GS-Doppler feature fusion network and the MULTITUDE framework. Detailed results are shown in Supplementary Table 7 and Supplementary Table 13.

The superiority of the GS-Doppler feature fusion network comes from the rich information from the two US modalities. On the one hand, Doppler US images provide information about synovial hypervascularity, which indicates the existence and position of synovial hypertrophy (Fig. 3a). On the other hand, GSUS images do not contain Doppler signals, thus are more suitable for AI models to focus on morphological characteristics of synovial hypertrophy (Fig. 3b). In addition, the superiority of the MULTITUDE framework comes from joint analysis of SH and vascularity score predictions from multiple AI models. Different from custom model ensemble strategy that ensembles multiple models separately for each classification tasks, MULTITUDE considers the relationship between the tasks (Fig. 3c).

**Fig. 3.**
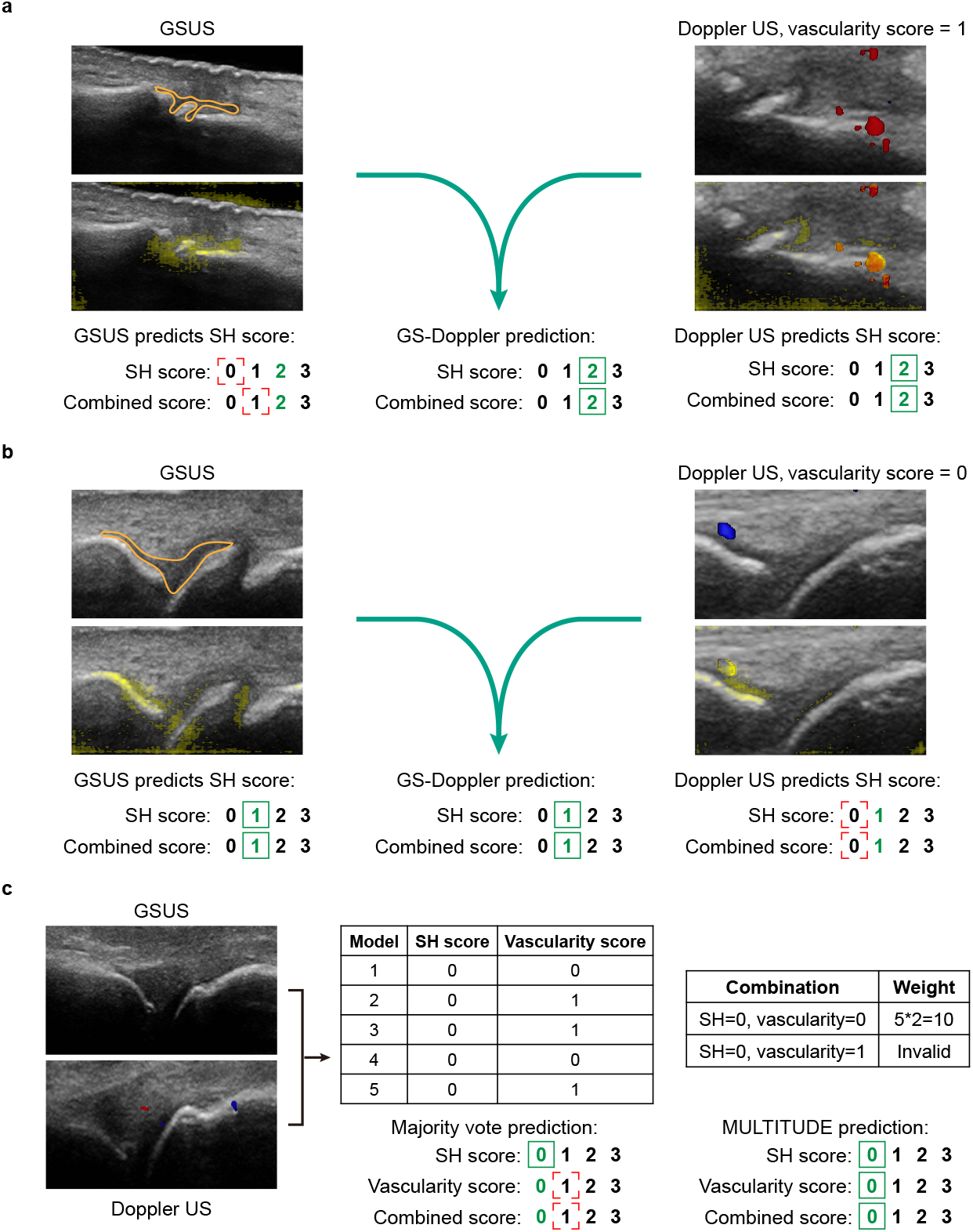
Superiority of the GS-Doppler feature fusion network and MULTITUDE. In each GSUS image, the synovial hypertrophy area is annotated in orange. Numbers in green are ground truth scores. Green rectangles in solid line stand for a correct prediction, while red rectangles in dashed line stand for a incorrect prediction. **a**, A sample of combined score grade 2 is underestimated as grade 1 using only the GSUS image. With the aid of synovial hypervascularity information in the Doppler US image, the RATING model makes the correct prediction. **b**, A sample of combined score grade 1 is underestimated as grade 0 using only the Doppler US image. With the aid of overall morphological changes of synovial hypertrophy in the GSUS image, the RATING model makes the correct prediction. **c**, A sample of combined score grade 0 is incorrectly predicted as grade 1 by custom majority vote method. MULTITUDE excludes the invalid score combination and makes the correct prediction.

### Explainability of the RATING model

Explainability of the RATING model is important to understand how it makes prediction and better assist radiologists. For each GSUS image or Doppler US image, the RATING model generate a heatmap that highlights the important areas for deciding the SH score. The heatmap of GSUS images may highlight the potential synovial hypertrophy area, and the heatmap of Doppler US images may highlight the potential synovial hypertrophy area and the potential synovial hypervascularity. The heatmaps are colorized in yellow and overlaid on the original US images. We found that the RATING model learnt the features of synovial hypertrophy and blood flow (Fig. 4).

**Fig. 4.**
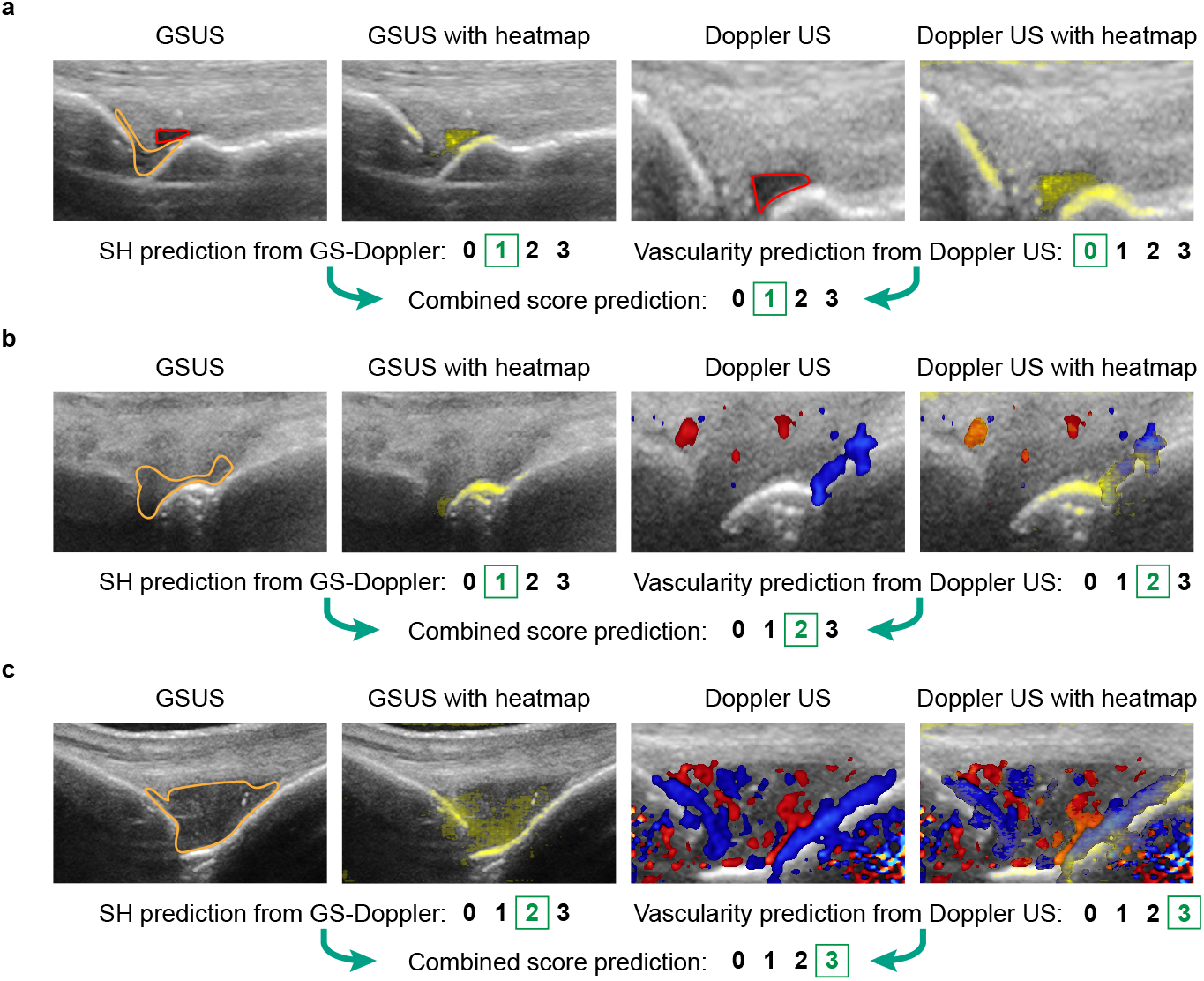
Examples of heatmap visualization. In each GSUS image, the synovial hypertrophy area is annotated in orange, and the joint effusion area is annotated in red. In each heatmap-overlaid image, the heatmap is colorized in yellow and overlaid on the original US image. **a**, A sample whose SH score is 1, vascularity score is 0, and combined score is 1. The joint effusion areas are highlighted in both the heatmaps of the GSUS and Doppler US image. **b**, A sample whose SH score is 1, vascularity score is 2, and combined score is 2. The synovial hypertrophy area near the bone surface is highlighted in the GSUS image and the Doppler US image, and the blood flow areas are highlighted in the Doppler US image. **c**, A sample whose SH score is 2, vascularity score is 3, and combined score is 3. The synovial hypertrophy area is highlighted in the GSUS image, and the blood flow areas are highlighted in the Doppler US image. The heatmaps show where the RATING model pays attention to, which human radiologists can use to understand the justification of the model for its prediction.

### Assistance of the RATING model to radiologists

To better assist radiologists for scoring RA, we developed a graphical user interface (Fig. 5) which displays the original US images, heatmap overlay images and AI predictions.

**Fig. 5.**
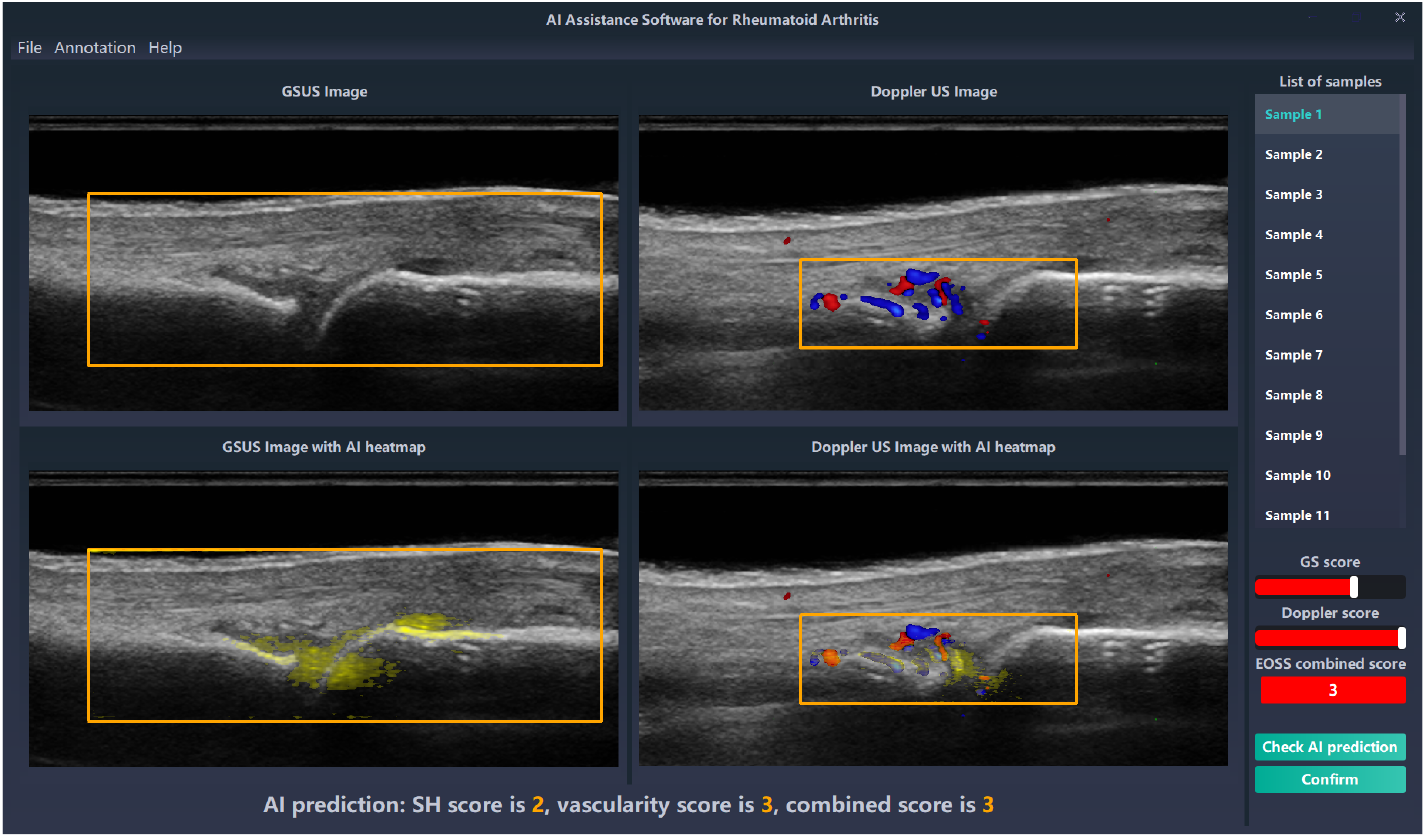
The graphical user interface of the RATING model to assist radiologists for scoring RA. The paired GSUS and Doppler US images are shown in the first row, and the ROI of each image is annotated by an orange rectangle. When radiologists click the button to check the RATING model’s outputs, the heatmap overlay images will be shown in the second row, and the AI predictions will appear at the bottom.

To evaluate the usefulness of the RATING model in assisting clinical decision making, we conducted a reader study and an AI-assisted reader study on the prospective test dataset. We recruited ten radiologists from PUMCH whose US experience range from 4 to 15 years. Detailed experience conditions are shown in Supplementary Table 17. We first conducted the reader study that radiologists independently scored the paired GSUS and Doppler US images in the prospective test dataset. We evaluated the performance of the RATING model and radiologists in two ways. Firstly, we calculated the four-class accuracy and linearly weighted *κ* for combined score classification. Secondly, we calculated the Youden index [20] in three combined score binary classification settings (that is, 0 versus 1,2 and 3, 0 and 1 versus 2 and 3, 0,1 and 2 versus 3). Besides the ten human readers, we compared the RATING model with an average reader for each evaluation metric. The RATING model achieved significantly higher combined score accuracy than the ten radiologists and the average reader (*P <* 0.001, Fig. 6a). In all the three combined score binary classification settings, the RATING model achieved significantly higher Youden index than the ten radiologists and the average reader (*P <* 0.001, Fig. 6b,c,d).

**Fig. 6.**
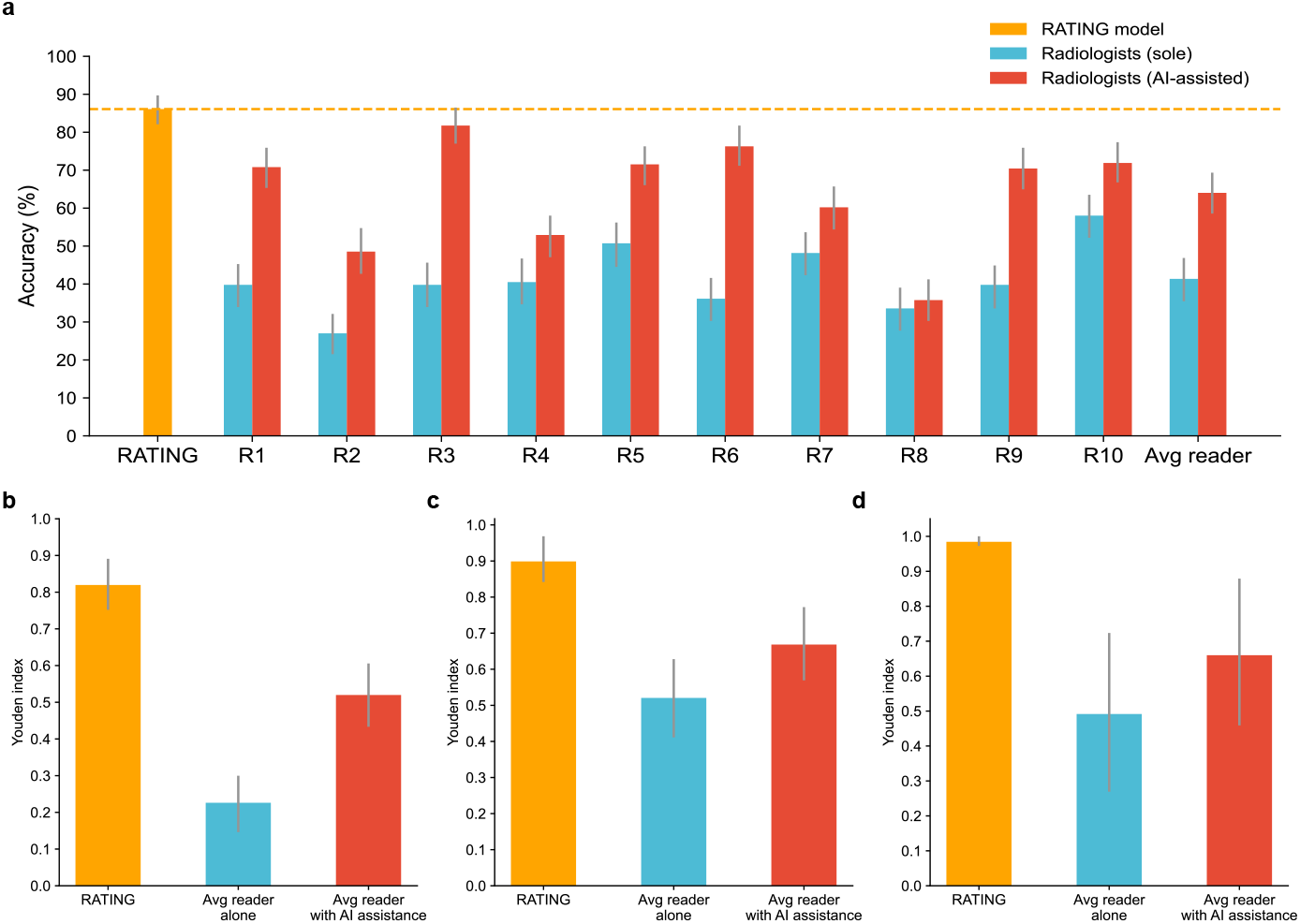
Performance comparison of the RATING model, radiologists alone and with AI assistance. **a**, With the assistance of the RATING model, radiologists (R1-R10) and the average reader achieved higher accuracy in the classification of combined score. **b-d**, The Youden index of radiologists’ combined score binary classification without and with the RATING model assistance: 0 versus 1,2 and 3 (**b**), 0 and 1 versus 2 and 3 (**c**), 0,1 and 2 versus 3 (**d**). Error bars indicate 95% confidence intervals.

After one week, we conducted the AI-assisted reader study that the same group of radiologists scored the same set of images with the assistance of the RATING model. With the assistance of the RATING model, the accuracy of the average reader was significantly improved from 41.4% (95% CI=35.8%– 47.2%) to 64.0% (95% CI=58.7%–69.5%), and all the ten radiologists achieved significantly higher combined score accuracy than independent assessment with *P <* 0.001 (Fig. 6a). Detailed accuracy results of the SH, vascularity and combined score are shown in Supplementary Table 18). The Youden index of the average reader was significantly improved from 0.226 to 0.520 on the classification of combined score in 0 versus 1,2 and 3 (*P <* 0.001), from 0.520 to 0.668 on the classification of combined score in 0 versus 1,2 and 3 (*P <* 0.001), and from 0.492 to 0.660 on the classification of combined score in 0 versus 1,2 and 3 (*P <* 0.001), as illustrated in Fig. 6b,c,d. Detailed Youden index results of all the ten radiologists and the average reader are shown in Supplementary Table 19.

## Discussion

With this work, we developed the RATING model to automatically assess US images for RA evaluation. In practice, the US quantitative assessment of disease activity is hampered by low intra-observer and inter-observer agreement during US examination. Moreover, it takes considerable time and expense to train experienced radiologists in US diagnostics. The RATING model is designed to assist radiologists in clinical practice, which not only provides all three score predictions instead of just the combined score prediction, but also generates heatmaps to indicate the areas that the AI models focus on to make predictions. We also developed a graphical user interface for radiologists, and conducted a reader study and an AI-assisted reader study to evaluate the effectiveness of its assistance to the radiologists in clinical trial settings. Experiment results demonstrated that the RATING model has great potential in improving the intra-observer and inter-observer agreement for radiologists of various US examination experience.

Previous studies did not evaluate the performance of AI models in the prospective setting, thus unable to fully prove the effectiveness of AI models in clinical practice. Therefore, we prospectively collected test dataset after the collection of development dataset, then evaluated the RATING model, conducted a reader study and an AI-assisted reader study on the prospective test dataset. The RATING model achieved high accuracy and linearly weighted *κ* in predicting the combined score, which also significantly outperformed the ten radiologists.

To demonstrate the generalizability of the RATING model, it should be evaluated on US images collected from external medical centers. Both the development dataset and the prospective test dataset were collected at PUMCH, in which all the Doppler US images were in CDUS mode. Apart from CDUS, PDUS is another commonly used Doppler US mode in RA assessment. To assess the generalizability, we evaluated the RATING model on the external test dataset collected at SZPH that consists of power Doppler US images. On the external test dataset, the RATING model achieved a comparable accuracy score of 85.0% (95% CI=80.5%–89.1%) compared to the accuracy score of 86.1% (95% CI=82.5%–90.1%) on the prospective test dataset, and achieved a comparable linearly weighted *κ* score of 0.853 (95% CI=0.806–0.900) compared to the linearly weighted *κ* score of 0.857 (95% CI=0.817–0.897) on the prospective test dataset. The results demonstrate that the RATING model generalizes well to different US operators and Doppler US modes.

Though the GS-Doppler feature fusion network achieves the best performance in predicting the SH score, it does not show superiority in predicting the vascularity score. The GSUS images contain no information about synovial hypervascularity, and radiologists only examine the Doppler US images to assess the synovial hypervascularity condition and decide the vascularity score in clinical practice. Furthermore, it also increases the over-fitting risk of the AI models. Experiment results also demonstrated that using the GS-Doppler feature fusion network obtained lower accuracy and linearly weighted *κ* in classifying the vascularity score.

It should be noted that the methods used in the RATING model can easily be extended to other medical examination tasks. The core ideas of GS-Doppler feature fusion network are the training of feature extraction networks for each imaging mode and the fusion of all imaging mode features. For multimodal US evaluations such as assessment of breast cancer and thyroid cancer, a feature extraction network can be built for each imaging mode and their features can be fused in the similar way as GS-Doppler feature fusion network did. Besides, MULTITUDE jointly considers the suggestions of the sub-tasks from multiple machine radiologists, and explores the correlations among them to obtain a more convincing radiological decision. This method is also appropriate for other medical diagnosis in which the radiological decision consists of multiple components, such as the TNM staging system [21] for malignant tumor classification which includes primary tumor (T), regional lymph node (N) and distant metastasis (M).

To better understand when and why the RATING model might make incorrect predictions, we analyzed the incorrect cases in the prospective test dataset and external test dataset. Based on the normalized confusion matrices in Fig. 2a and Fig. 2c, we analyzed the four most common types of incorrect predictions and chose one typical example for each type (Fig. 7). Incorrect predictions may occur when the situation is just on the borderline between two grades, such as grade 0 and 1, grade 1 and 2, as well as grade 2 and 3. When this happens, human experts should carefully analyze the US images according to the EOSS guidelines.

**Fig. 7.**
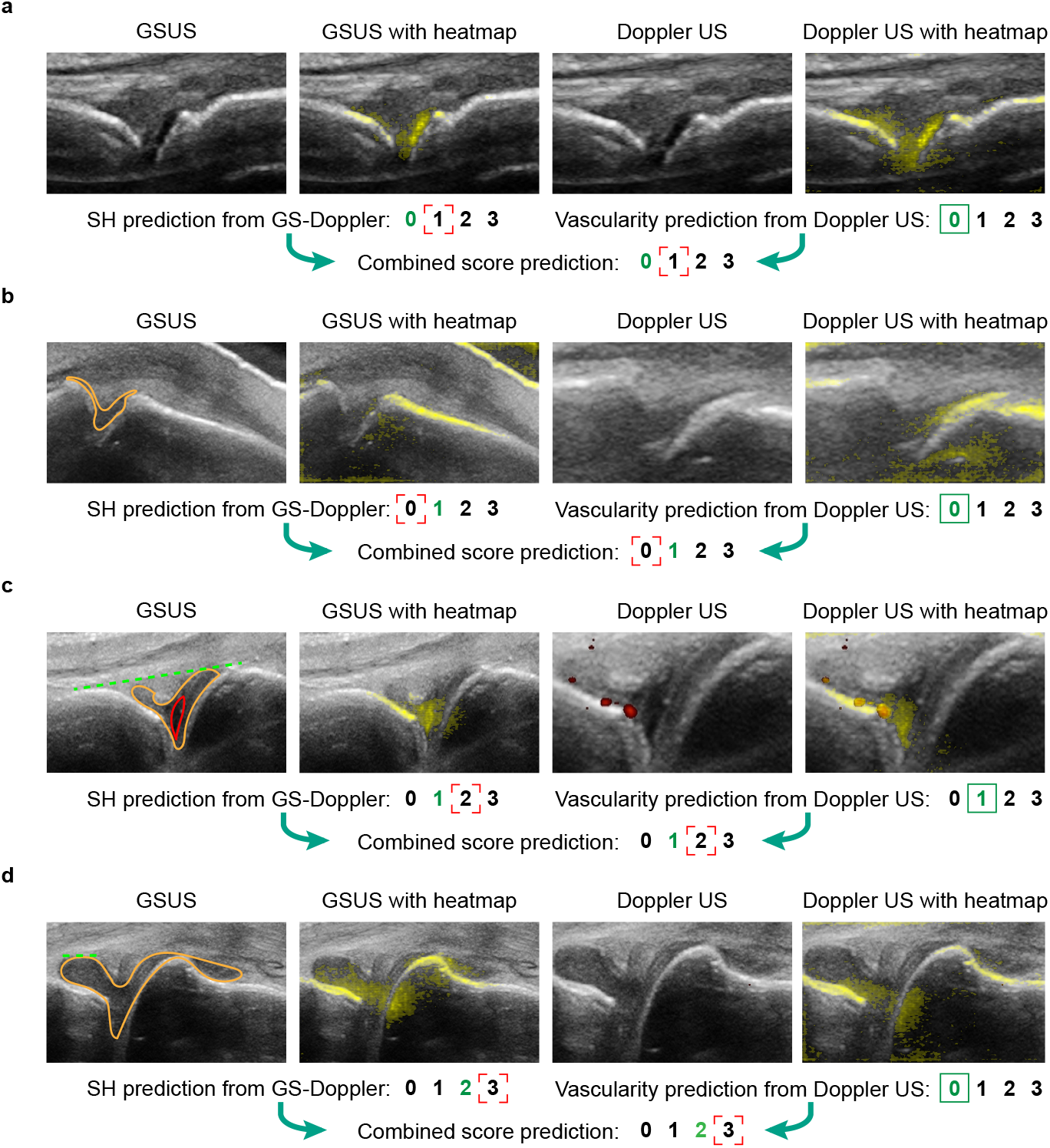
Typical examples of incorrect predictions. In each GSUS image, the synovial hypertrophy area is annotated in orange, and the joint effusion area is annotated in red. In each heatmap overlay image, the heatmap is colorized in yellow and overlaid on the original US image. Numbers in green are ground truth scores. Green rectangles in solid line stand for a correct prediction, while red rectangles in dashed line stand for a incorrect prediction. **a**, The sample of grade 0 is incorrectly predicted as grade 1. The model correctly identifies the mild synovial hypertrophy in both GSUS and Doppler US images, but overestimates it and predicts the combined score as 1. **b**, The model underestimates the mild synovial hypertrophy and incorrectly predicts the sample of grade 1 as grade 0. **c**, The sample of grade 1 is incorrectly predicted as grade 2, which may be resulted from the joint effusion area. Although there is obvious synovial hypertrophy and effusion, their areas do not exceed the joint line across left and right bones which is illustrated by the green dashed line. **d**, The sample of grade 2 is incorrectly predicted as grade 3. The SH score is determined by expert radiologists as 2 rather than 3 because the surface of the left synovial hypertrophy area is only slightly convex rather than obviously convex, which is just on the borderline between grade 2 and 3. The green dashed line illustrates the synovial hypertrophy surface line which is approximately horizontal.

One limitation of our study is about the test data. Because we collected data from two hospitals in China, generalizability of the RATING model to other population has not been demonstrated. All US images were acquired using Mindray US machines, therefore generalizability of the RATING model to US machines from other manufacturers has not been demonstrated. Future evaluation of the RATING model should include data from other population and other US machine manufacturers.

Another potential limitation of our study is that the RATING model consists of 60 convolutional neural networks, which might consume considerable inference time and result in applicability issues. Since these networks are independent to each other, one possible solution is to infer the input US images in parallel using multi-processing technique.

In conclusion, we propose the RATING model to evaluate the RA activity from GSUS and Doppler US images. We demonstrate that the RATING model outperforms all existing methods and implements well in clinical trial settings. Moreover, we demonstrate that RATING generalizes well to different US operators and Doppler US modes. The generated heatmaps make the RATING model more understandable, and the RATING model show great potential in assisting radiologists. In the future, it is promising to incorporate the RATING model into the US machines, displaying heatmaps and AI predictions on the screen in real time. Besides the clinical assistance, the RATING model may also be able to train junior radiologists.

## Methods

### Ethical approval

The study was divided into two parts. The model construction was based on the retrospective data collection, and the validation process was built prospectively. The study was registered at ClinicalTrial.gov (NCT04297475) and approved by the Institutional Review Boards of Peking Union Medical College Hospital (Approval number: JS-1923). The prospective study was a observational one and did not involve interventional methods. The recruited patients of the retrospective and prospective parts were well informed of the study and provided signed inform consent. Patients or public are not involved in design, recruitment and conduct of the study.

### Patients and data collection

To build the development dataset, we retrospectively collected GSUS and Doppler US images from patients who were diagnosed with RA according to the 2010 American College of Rheumatology/European League Against Rheumatism (ACR/EULAR) classification criteria between March 2019 to April 10th 2021 at the Peking Union Medical College Hospital (PUMCH). To evaluate performance of the RATING model, we prospectively recruited patients with RA between April 20th 2021 and October 2021 at PUMCH. To further evaluate the generalizability of the RATING model across Doppler modes, we recruited patients between March 2021 and December 2021 at Shenzhen People’s Hospital (SZPH). In retrospective, prospective and external workflow, the US images of the patients who were complicated with other inflammatory joint diseases were excluded. Details of data collection workflow are illustrated in Supplementary Fig. 1.

A total of four US operators from PUMCH and Shenzhen People’s Hospital performed US scanning. Two US operators from PUMCH both have 5 years of experience in musculoskeletal ultrasound (MSK-US) and about 1,000 MSK-US scanning cases per year. The other two US operators from Shenzhen People’s Hospital have 4 and 5 years of experience in MSK-US and about 700 MSK-US scanning cases per year. All the four US operators received training programs of the standard scanning protocol for small joints. In both sites, the recruited patients received US scanning of metacarpophalangeal (MCP) joints and proximal interphalangeal (PIP) joints using the same commercial ultrasound system (Resona 7, Mindray Bio-Medical Electronics Co., Ltd.) and probe (L23-15MHz, centrual frequency of 20MHz, Mindray Bio-Medical Electronics Co., Ltd.). For each patient, GSUS and Doppler US images were performed consecutively at a depth of 1.5-2 cm. The CDUS settings included pulse repetition frequency (PRF) of 1000 Hz, wall filter of 80 Hz, maximum gain of 50, and scale of 3 cm/s. The PDUS settings included PRF of 700 Hz, wall filter of 37 Hz, scale of 3 cm/s, maximum gain of 50. And both have a rectangle sampling box with no angulation. During the examination, patients placed their hands on the white surface of examining tables with a bubble-free gel pat put on the dorsal side of the hands. The operator positions the probe longitudinally on the dorsal surface of the patient’s fingers. Static GS image showing the bone surfaces of the long bones at both ends and synovial area clearly were collected. Corresponding Doppler US images on the same section were also saved subsequently.

After image collection, three experienced radiologists in PUMCH selected and scored GSUS and Doppler images for further analysis. The three radiologists have 10, 7, and 6 years of experience in US and 5, 5, 4 years of experience in MSK-US. They also read over 1,000 sets of MSK-US images per year. And they have received a six-month training program about EOSS system before conducting the study. The exclusion criterion for the images included: (1) images with significant artifacts (GS: blurred images, anisotropic artifacts; Doppler US: aliasing, motion artifacts); (2) images not clearly showing bone surfaces and synovium. Then the radiologists scored the selected GSUS and Doppler US images according to the EOSS system [5]. After completing scoring of all images, the three radiologists discussed the images with inconsistent scores until achieving consensus. When disagreement still existed, another professional radiologist from PUMCH with 15 years of experience in US and 10 years of experience in MSK-US re-evaluated the images and made the final decision. All the participated radiologists and US operators were blind to the patients’ clinical information.

### Image preprocessing

To eliminate irrelevant information, for each GSUS image and Doppler US image, the ROI was annotated as a rectangular area by experienced radiologists using a custom annotation software. Then the ROIs of US images were cropped and resized to 224 × 224 pixel size. To support both CDUS and PDUS modes, each Doppler US image was first segmented to obtain a binary mask of colour area. The binary mask had the same size as the original Doppler US image and pixel values were either 0 or 1, where 1 indicated the detection of blood flow. Afterwards, a masked Doppler US image was generated for each Doppler US image by assigning red colour to pixels in the original image where the corresponding pixel in the binary mask was 1.

### Overall pipeline of RATING

Based on the development dataset, we proposed the RATING model (Supplementary Fig. 2) which mimics the process that radiologists discuss and jointly consider SH and vascularity score to reach agreement on RA scoring. During inference stage, for each sample, five models separately predict the SH score and the vascularity score, and the combined score is predicted by comprehensively considering five models’ predictions using our proposed MULTI-Task mUlti-moDel Ensemble method named MULTITUDE. Model ensemble is a type of machine learning technique that combines a number of weak learners to achieve better performance than each individual learners. Custom model ensemble methods such as stacking [22] and AdaBoost [23] solve only one prediction task at a time. Recently, the idea of ensemble has been introduced to multi-task learning [24], but each task is predicted separately. Our proposed MULTITUDE jointly considers multiple classification tasks and utilizes restrictions on value combinations. To enhance the accuracy and robustness of the RATING model, the five models were expected to learn diverse features under different training settings. We randomly partitioned the development dataset into five complementary subsets of an equivalent number of samples, and every four of the five subsets were used to train a model where the remaining subset was used to validate the model. Therefore, a total of five models were obtained, mimicking five radiologists who were trained on different data.

Each of the five models is composed of a SH scoring module that predicts the SH score and a vascularity scoring module that predicts the vascularity score, which both are four-class classification tasks. Instead of directly predicting the scores by multi-classification models, we adopted a technique called error-correcting output codes (ECOC) [25] that solves the four-class classification tasks by a series of binary classification models. Specifically, SH scoring modules and vascularity scoring modules share the same strategy to solve four-class classification tasks by three binary classification tasks, i.e., whether the score was greater than 0, 1 and 2. Theoretically, at least three binary classification models are needed to solve a four-class classification task. To improve accuracy and robustness, for each binary classification task, we trained two networks with the same training settings except different randomization seed zero and one, resulting in a total of six classification networks. With the six binary classification predictions, the four-class classification prediction was calculated using ECOC.

For binary classification networks of SH scoring modules, we propose GS-Doppler feature fusion network that jointly analyzes GSUS and Doppler US images. The GS-Doppler feature fusion network mimics the practical US assessment procedure that radiologists assess both GSUS and Doppler US images to decide the SH score. To utilize features from multimodal US images, previous studies [9] simply extract features using networks pre-trained on ImageNet [26]. Instead, our proposed GS-Doppler feature fusion network is first pre-trained with self supervision and then fine-tuned on the target classification task, thus is capable of extracting more appropriate features for the target task. For binary classification networks of vascularity scoring modules, instead of using the GS-Doppler feature fusion network, we only used the Doppler US images to make predictions. This is because radiologists only examine the Doppler US images to assess the synovial hypervascularity condition and decide the vascularity score in clinical practice.

### MULTITUDE

The propose of MULTITUDE (Supplementary Fig. 2a) is inspired by the process that radiologists discuss and jointly consider SH and vascularity score to reach agreement on RA scoring. However, the usage of MULTITUDE is not limited to RA assessment. Generally, we define a medical diagnosis problem where a total of *t* measurements *S*_1_, *S*_2_, …, *S*_*t*_ need to be classified. For any measurement *S*_*i*_ (*i* = 1, 2, …, *t*), its all *c*_*i*_ possible values are denoted as 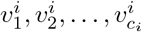. For the above problem, *m* models ℳ_1_, ℳ_2_, …, ℳ_*n*_ are built to mimic *m* independent radiologists. For any model ℳ_*j*_ (*j* = 1, 2, …, *m*), it separately predicts the *k* measurements as 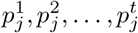 where 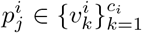. We define 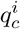 as the number of models that predicts *S*_*i*_ (*i* = 1, 2, …, *t*) as 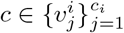. It should satisfy equation 1:

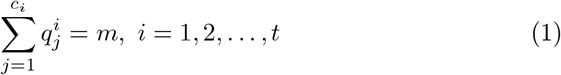

We use the term *value combination* to refer a possible condition of the *t* measurements, which is represented as a tuple (*v*^1^, *v*^2^, …, *v*^*t*^). Theoretically, there are 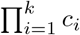 possible value combinations of the *t* measurements. However, some value combinations contradict with medical knowledge. MULTITUDE figures out all valid combinations and calculate the weight for each valid combination. The weight of a valid combination (*v*^1^, *v*^2^, …, *v*^*t*^) quantifies the agreement of the models on it, which is defined as 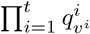. Larger weight of a valid combination indicates that more models agree on it. Therefore, the final predictions of the *t* measurements are obtained from the valid combination with the largest weight. If more than one valid combination gains the largest weight, the first one in alphabetical order is selected.

Different from the custom majority vote ensemble strategy which separately makes prediction for each measurement, MULTITUDE takes advantage of relationships between different measurements. As a result, invalid value combinations are excluded by MULTITUDE, leading to higher classification accuracy. If all value combinations are valid, MULTITUDE will yield the same prediction as the majority vote ensemble.

As for RA assessment, the combined score can be obtained directly from the SH score and the vascularity score, thus only the latter two measurements need to be predicted. Both the SH score and the vascularity score range from 0 to 3, resulting in 16 theoretically possible combinations. According to the EOSS system, three combinations are invalid whose SH score is 0 and vascularity score is greater than 0, so only 13 combinations are valid. We used *m* = 5 models in this paper.

### GS-Doppler feature fusion network

GS-Doppler feature fusion network (Supplementary Fig. 2c) comprehensively analyzes a sample’s GSUS and Doppler US image to predict SH score binary classification probability. It is composed of a GSUS feature extraction network ℱ, a Doppler US feature extraction network 𝒢 and a fusion classification network ℋ. ℱ and 𝒢 are ResNet-18 [27] networks that extract high-level feature vectors from GSUS images and Doppler US images. ℋ is a two-layer multi-layer-perceptron (MLP) that fuses the high-level feature vectors from the GSUS image and the Doppler US image of a sample and predicts the SH score.

During inference time, for a sample of a GSUS ROI *x*_G_ and a masked Doppler US ROI *x*_D_, ℱ extracts a 512-dimension feature vector *h*_G_ from *x*_G_, and 𝒢 extracts a 512-dimension feature vector *h*_D_ from *x*_D_. *h*_G_ and *h*_D_ contains potential useful information in the GSUS image and the Doppler US image for predicting the SH score. Subsequently, *h*_G_ and *h*_D_ are fused by concatenating them into a 1024-dimension feature vector *h*_fusion_ and feeding it into ℋ. The GS-Doppler feature fusion network is trained in a 3-stage manner. In the first stage, a technique called self-supervised pre-training is adopted to train a ResNet-18 network that learns both a feature mapping of joint parts in US images as well as their correct spatial arrangement [28]. To be specific, each ROI of GSUS image is resized to 225 × 225 and split into a 3 × 3 grid, then a 64 × 64 tile is randomly cropped from each 75 × 75 grid cell. These 9 tiles are re-ordered via a randomly chosen permutation from 1000 predefined permutations and then fed to the ResNet-18 network to obtain 9 feature vectors. Finally, these feature vectors are concatenated and fed into a MLP to predict probabilities that the chosen permutation belongs to 1000 predefined permutations. In the second stage, the pre-trained ResNet-18 network in the first stage is maintained as initial weights and further trained to extract features that contain potential useful information in GSUS and Doppler US images for predicting the SH score. Thus, ℱ and 𝒢 are obtained by training for the SH score binary classification task using GSUS images and masked Doppler US images, respectively. In the third stage, with ℱ and 𝒢 already obtained, ℋ is trained for the SH score binary classification task.

### Training details

We implemented our models on the PyTorch deeplearning framework. Cross entropy loss was used to optimize classification networks. All networks were optimized using an adaptive moment estimation (ADAM) optimizer [29] in a batch size of 64 with an initial learning rate of 0.0003, which then decayed every 15 epochs with a decay factor of 0.3. To address the class imbalance issue, images are randomly re-sampled at the beginning of every epoch so that there were same amount of training samples in different classes. Data augmentation was also performed that images in training set were augmented by applying random cropping and color jittering. To aid training of neural networks, we adopted transfer learning strategy that the ResNet-18 network pre-trained on ImageNet [26] was used as initial weights for feature extraction networks and vascularity score classification networks.

### Heatmap generation

To assure trust by human experts and assist radiologists in clinical setting, heatmaps were calculated from GS-Doppler feature fusion networks that predict whether the SH score was greater than 0, indicating potential synovial hypertrophy area. The integrated gradient (IG) [30] technique was recently proposed that assigns an importance score to each input feature. For a model ℳ(·) and an input *x*, we defined the baseline input *x*′ as a zero-filled tensor that has the same shape as *x*, and defined integrated gradient as the path integral of the gradients along the straight-line path from *x*′ to *x*. Specifically, for an input *x* of *n* features 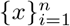, the integrated gradient along *x*_*i*_ was defined using equation 2:

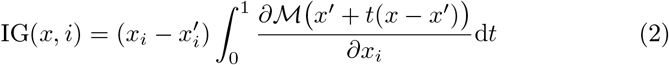

In practice, the integration was approximated via a summation using equation 3:

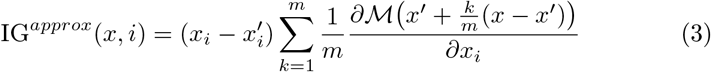

where we used *m* = 50 in all experiments. Thus *n* integrated gradients were obtained, and they formed a new image 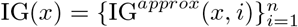 that was of the same size as the input image *x*.

To make the calculation of heatmaps more robust, Gaussian noise *δ* ∼ *𝒩* (0, 1) was randomly added to the original image *x* for eight times, and eight heatmaps were generated using IG. The final heatmap for model ℳ and input *x* was generated by averaging the eight heatmaps. To visualize the heatmaps, a yellow mask was overlaid on the original image *x* of which alpha channel pixel values were decided by corresponding heatmap pixel values. To remove less important features, pixel values smaller than 0.2 are set to zero.

In the RATING model, two GS-Doppler feature fusion networks were trained that predict whether SH score was greater than 0 in each of the five models, thereby ten heatmaps were generated for each input *x*. Due to the difference of training data and randomization seed, knowledge learned by networks might also differ, resulting in different attention areas in the same input image. To combine knowledge of all the networks, we averaged all the heatmaps to generate a GSUS heatmap and a Doppler US heatmap, as shown in Supplementary Fig. 9.

### Reader study

A reader study was conducted to compare the performance of the RATING model with that of the ten radiologists, whose US examination experiences are shown in Supplementary Table 17. A total of 274 samples from 28 patients in the prospective test dataset were presented to radiologists and the RATING model in random order. For each sample, ROIs of GSUS and Doppler US were provided. The radiologists were blinded to each other and to the AI outputs. Each radiologist reviewed the same set of samples independently and decided the SH score, the vascularity score and the combined score according to the EOSS system.

### AI-assisted reader study

To evaluate the assistance of the RATING model to the radiologists in guiding clinical decisions, we conducted an AI-assisted reader study. The same 274 samples in the prospective test dataset were presented to the same ten radiologists again in a different order. For each sample, together with the ROIs of the GSUS and Doppler US image, the RATING model’s heatmaps and predictions on the SH score, the vascularity score and the combined score were provided to the readers at the same time. The radiologists were blind to the first-time interpretation and to each other. The heatmaps indicated potential synovial hypertrophy area and synovial hypervascularity area, and the scores predicted by the RATING model served as a reference which might assist the readers when they lacked confidence in their own judgement.

### Statistical analysis

For SH score binary classification tasks and vascularity score binary classification tasks where ten models were trained in each task, the 95% CI of AUC, PPV, NPV, sensitivity and specificity were computed as 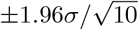 where *σ* was standard error across the ten models. For the ablation method for MULTITUDE and the baseline cascade method [17], the 95% CI were computed as 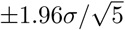 where *σ* was standard error across the five models. For the remaining experiments where only a single accuracy and a single *κ* value were available, we bootstrapped the estimation for 1,000 iterations and reported the 2.5th and 97.5th percentiles as the 95% CI.

The agreement between AI models and experienced radiologists was graded as follows: poor (*κ* ≤ 0.20), moderate (0.20 *< κ* ≤ 0.40), fair (0.40 *< κ* ≤ 0.60), good (0.60 *< κ* ≤ 0.80), or very good (0.80 *< κ* ≤ 1.00). To compare differences between accuracy scores and between linearly weighted *κ*, we bootstrapped the estimation for 1,000 iterations and compared using z-test. All statistical tests were two sided and *P* values *<* 0.001 indicated statistically significant differences. The analyses were performed using Python scikit-learn library and statsmodel library.

## Data Availability

All data produced in the present work are contained in the manuscript

## Data availability

The datasets generated during and/or analysed during the current study are not publicly available due to hospital regulations.

## Code availability

The codes for building and testing RATING are publicly available at http://39.102.45.122/zhouzhanping/rating.

## Author contributions

Z.Z. developed the system and analyzed experiment results. Z.Z. and C.Z. wrote the manuscript. C.Z., M.W. and M.Y. provided clinical expertise. Z.Z., H.Q. and Y.G. discussed the techniques. C.Z., M.W., Q.W. and R.Z. created the datasets and scored the US images. Q.W., R.Z., H.W., F.D., Z.Q., X.T. and X.Z. collected the data. J.L. and Y.J. discussed medical issues. H.Q, F.X. and M.Y. revised the manuscript. H.Q, F.X., Q.D. and M.Y. supervised the study.

## Competing interests

The authors declare no competing interests.

## Notes

### Competing Interest Statement

The authors have declared no competing interest.

### Clinical Trial

NCT04297475

### Funding Statement

This study was funded by National Natural Science Foundation of China
(Nos.: 61971447, 81301268, and 81421004), Beijing Natural Science Foundation
(No.: JQ18023), CAMS Innovation Fund for Medical Sciences (No.: 2020-I2M-
C&T-B-035), International Science and Technology Cooperation Programme
(No.: 2015DFA30440), Chinese National Key Technology R&;D Program, Min-
istry of Science and Technology (Nos.: 2019YFC0840603, 2017YFC0907601,
2017YFC0907604, and 2017YFE0104200), CAMS Innovation Fund for Medical
Sciences (No.: 2021-I2M-1-005), and The Non-profit Central Research Institute
Fund of Chinese Academy of Medical Sciences (No.: 2021-PT320-002).

### Author Declarations

The Institutional Review Boards of Peking Union Medical College Hospital gave ethical approval for this work

