## Supplementary Information for "RATING: Medical-knowledge-guided rheumatoid arthritis assessment from multimodal ultrasound images via deep learning"

### Supplementary Material

16 **Supplementary Table 16: Confusion matrix of EOSS combined score prediction on the external test dataset** 11

| | | |
|---|---|---|
| 17 | **Supplementary Table 17: Experience of radiologists (R1-R10) in reader study and AI-assisted reader study** | 12 |
| 18 | **Supplementary Table 18: Accuracy of radiologists (R1-R10) and the average reader in reader study and AI-assisted reader study** | 13 |
| 19 | **Supplementary Table 19: Youden index of radiologists (R1-R10) and the average reader in reader study and AI-assisted reader study** | 14 |
| 1 | **Supplementary Figure 1: Overview of the retrospective, prospective and external study workflow** | 15 |
| 2 | **Supplementary Figure 2: Structure of the RATING model.** | 16 |
| 3 | **Supplementary Figure 3: Performance of three SH score binary classification tasks on the development dataset** | 17 |
| 4 | **Supplementary Figure 4: Performance of three vascularity score binary classification tasks on the development dataset** | 17 |
| 5 | **Supplementary Figure 5: Performance of three SH score binary classification tasks on the prospective test dataset** | 18 |
| 6 | **Supplementary Figure 6: Performance of three vascularity score binary classification tasks on the prospective test dataset** | 18 |
| 7 | **Supplementary Figure 7: Performance of three SH score binary classification tasks on the external test dataset** | 19 |
| 8 | **Supplementary Figure 8: Performance of three vascularity score binary classification tasks on the external test dataset** | 19 |
| 9 | **Supplementary Figure 9: Illustration of heatmap overlay image generation process** | 20 |

**Supplementary Table 1. EOSS system guidelines.**

| Category | | Definition |
|---|---|---|
| SH score | Grade 0 | No SH independently of the presence of effusion. |
| | Grade 1 | SH with or without effusion up to level of horizontal line connecting bone surfaces metacarpal head and proximal phalangeal bone. |
| | Grade 2 | SH with or without effusion extending beyond joint line but with upper surface convex (curved downwards) or hypertrophy extending beyond joint line but with upper surface flat. |
| | Grade 3 | SH with or without effusion extending beyond joint line but with upper surface flat or convex (curved downwards). |
| Vascularity score | Grade 0 | No Doppler activity. |
| | Grade 1 | Up to three single Doppler spots or up to one confluent spot and two single spots or up to two confluent spots. |
| | Grade 2 | Greater than Grade 1 but $< 50\%$ Doppler signals in the total GS background. |
| | Grade 3 | Greater than Grade 2 ($> 50\%$ of the background GS). |
| Combined score | Grade 0 | No GS-detected SH and no Doppler signal (within the synovium). |
| | Grade 1 | Grade 1 SH and $\leq$ Grade 1 Doppler signal. |
| | Grade 2 | Grade 2 SH and $\leq$ Grade 2 Doppler signal or Grade 1 SH and a Grade 2 Doppler signal. |
| | Grade 3 | Grade 3 SH and $\leq$ Grade 3 Doppler signal or Grade 1 or 2 SH and a Grade 3 Doppler signal. |

**Supplementary Table 2. Patient demographics and sample characteristics.**

| Characteristics | Development dataset | Prospective test dataset | External test dataset |
|---|---|---|---|
| Number of patients | 104 | 28 | 42 |
| Mean age (years) | 54.6 (23-80) | 44.7 (25-70) | 46.9 (28-80) |
| Gender | | | |
| Male | 25 (24.0%) | 14 (50.0%) | 13 (31.0%) |
| Female | 79 (76.0%) | 14 (50.0%) | 29 (69.0%) |
| Number of samples | 752 | 274 | 293 |
| Joint type | | | |
| PIP | 333 (44.3%) | 132 (48.2%) | 86 (29.4%) |
| MCP | 419 (55.7%) | 142 (51.8%) | 207 (70.6%) |
| SH score | | | |
| 0 | 310 (41.2%) | 152 (55.5%) | 174 (59.4%) |
| 1 | 188 (25.0%) | 73 (26.6%) | 56 (19.1%) |
| 2 | 181 (24.1%) | 43 (15.7%) | 38 (13.0%) |
| 3 | 73 (9.7%) | 6 (2.2%) | 25 (8.5%) |
| Vascularity score | | | |
| 0 | 519 (69.0%) | 198 (72.3%) | 214 (73.0%) |
| 1 | 130 (17.3%) | 31 (11.3%) | 23 (7.8%) |
| 2 | 72 (9.6%) | 31 (11.3%) | 29 (9.9%) |
| 3 | 31 (4.1%) | 14 (5.1%) | 27 (9.2%) |
| Combined score | | | |
| 0 | 296 (39.4%) | 148 (54.0%) | 173 (59.0%) |
| 1 | 188 (25.0%) | 55 (20.1%) | 54 (18.4%) |
| 2 | 179 (23.8%) | 53 (19.3%) | 34 (11.6%) |
| 3 | 89 (11.8%) | 18 (6.6%) | 32 (10.9%) |

**Supplementary Table 3. Performance of three SH score binary classification tasks on the development dataset.** Data in parentheses are 95% confidence intervals.

| Task | AUC | PPV | NPV | Sensitivity (%) | Specificity (%) |
|---|---|---|---|---|---|
| 0 vs 1,2,3 | 0.896(0.883-0.909) | 0.869(0.858-0.880) | 0.773(0.739-0.808) | 82.7(79.1-86.2) | 82.3(80.6-84.0) |
| 0,1 vs 2,3 | 0.945(0.935-0.956) | 0.782(0.748-0.817) | 0.939(0.927-0.951) | 88.8(86.4-91.1) | 87.1(84.5-89.6) |
| 0,1,2 vs 3 | 0.948(0.932-0.964) | 0.453(0.420-0.486) | 0.990(0.984-0.995) | 92.4(88.6-96.2) | 87.7(85.8-89.7) |

**Supplementary Table 4. Performance of three vascularity score binary classification tasks on the development dataset.** Data in parentheses are 95% confidence intervals.

| Task | AUC | PPV | NPV | Sensitivity (%) | Specificity (%) |
|---|---|---|---|---|---|
| 0 vs 1,2,3 | 0.980(0.976-0.984) | 0.871(0.848-0.894) | 0.969(0.959-0.979) | 93.4(91.7-95.2) | 93.6(92.1-95.0) |
| 0,1 vs 2,3 | 0.992(0.988-0.996) | 0.795(0.705-0.884) | 0.998(0.996-1.000) | 98.9(97.6-100.3) | 95.4(93.4-97.5) |
| 0,1,2 vs 3 | 0.991(0.986-0.995) | 0.656(0.531-0.781) | 1.000(1.000-1.000) | 100.0(100.0-100.0) | 96.9(95.3-98.5) |

**Supplementary Table 5. Performance of three SH score binary classification tasks on the prospective test dataset.** Data in parentheses are 95% confidence intervals.

| Task | AUC | PPV | NPV | Sensitivity (%) | Specificity (%) |
|---|---|---|---|---|---|
| 0 vs 1,2,3 | 0.930(0.919-0.941) | 0.846(0.822-0.871) | 0.885(0.879-0.890) | 85.8(84.9-86.7) | 87.3(84.8-89.8) |
| 0,1 vs 2,3 | 0.933(0.930-0.936) | 0.577(0.552-0.602) | 0.974(0.971-0.978) | 89.6(88.0-91.2) | 85.5(83.9-87.1) |
| 0,1,2 vs 3 | 0.979(0.973-0.985) | 0.358(0.281-0.435) | 1.000(1.000-1.000) | 100.0(100.0-100.0) | 95.3(93.8-96.8) |

**Supplementary Table 6. Performance of three vascularity score binary classification tasks on the prospective test dataset.** Data in parentheses are 95% confidence intervals.

| Task | AUC | PPV | NPV | Sensitivity (%) | Specificity (%) |
|---|---|---|---|---|---|
| 0 vs 1,2,3 | 0.986(0.985-0.987) | 0.877(0.863-0.890) | 0.986(0.982-0.990) | 96.2(95.1-97.4) | 95.1(94.5-95.8) |
| 0,1 vs 2,3 | 0.990(0.986-0.995) | 0.805(0.773-0.837) | 0.994(0.991-0.997) | 97.0(95.6-98.5) | 95.4(94.5-96.3) |
| 0,1,2 vs 3 | 0.995(0.991-0.998) | 0.736(0.658-0.813) | 1.000(1.000-1.000) | 100.0(100.0-100.0) | 97.9(97.1-98.6) |

**Supplementary Table 7. Accuracy and linearly weighted $\kappa$ comparison on the prospective test dataset.** Data in parentheses are 95% confidence intervals. $P$ values indicate comparison between RATING model and other methods.

| Metric | SH score | Vascularity score | Combined score | $P$ value |
|---|---|---|---|---|
| Accuracy(%) | | | | |
| RATING model | 79.6(74.8,84.3) | 94.5(91.6,97.1) | 86.1(82.5,90.1) | - |
| SH prediction using GSUS | 73.7(68.6,79.2) | 94.2(91.2,96.7) | 80.3(75.5,85.0) | $< 0.001$ |
| SH prediction using Doppler US | 73.4(67.5,78.8) | 94.2(91.2,96.7) | 79.9(75.2,85.0) | $< 0.001$ |
| Vascularity prediction using both modes | 79.6(74.8,84.3) | 92.7(89.8,95.6) | 84.7(80.7,89.1) | $< 0.001$ |
| w/o MULTITUDE | 73.9(72.7-75.0) | 92.6(91.8-93.4) | 80.1(78.9-81.4) | $< 0.001$ |
| Multi-classification | 69.1(67.9-70.2) | 88.0(86.2-89.7) | 72.5(70.7-74.3) | $< 0.001$ |
| Cascade [1] | - | - | 69.0(66.7-71.2) | $< 0.001$ |
| Linearly weighted $\kappa$ | | | | |
| RATING model | 0.757(0.699-0.885) | 0.919(0.876-0.966) | 0.853(0.806-0.900) | - |
| SH prediction using GSUS | 0.674(0.607-0.869) | 0.914(0.870-0.966) | 0.792(0.738-0.847) | $< 0.001$ |
| SH prediction using Doppler US | 0.685(0.623-0.764) | 0.914(0.870-0.966) | 0.795(0.742-0.847) | $< 0.001$ |
| Vascularity prediction using both modes | 0.757(0.699-0.814) | 0.903(0.862-0.943) | 0.841(0.794-0.887) | $< 0.001$ |
| w/o MULTITUDE | 0.689(0.681-0.698) | 0.897(0.887-0.908) | 0.792(0.779-0.805) | $< 0.001$ |
| Multi-classification | 0.606(0.587-0.626) | 0.811(0.779-0.843) | 0.698(0.674-0.721) | $< 0.001$ |
| Cascade [1] | - | - | 0.608(0.569-0.647) | $< 0.001$ |

**Supplementary Table 8. Confusion matrix of SH score prediction on the prospective test dataset.**

| True label | Model prediction | | | | Total | Accuracy (%) |
|---|---|---|---|---|---|---|
| | 0 | 1 | 2 | 3 | | |
| 0 | 142 | 9 | 1 | 0 | 152 | 93.4 |
| 1 | 12 | 40 | 21 | 0 | 73 | 54.8 |
| 2 | 2 | 5 | 30 | 6 | 43 | 69.8 |
| 3 | 0 | 0 | 0 | 6 | 6 | 100.0 |
| Total | 152 | 73 | 43 | 6 | 274 | 79.6 |

**Supplementary Table 9. Confusion matrix of vascularity score prediction on the prospective test dataset.**

| True label | Model prediction | | | | Total | Accuracy (%) |
|---|---|---|---|---|---|---|
| | 0 | 1 | 2 | 3 | | |
| 0 | 197 | 3 | 2 | 0 | 202 | 97.5 |
| 1 | 4 | 20 | 4 | 0 | 28 | 71.4 |
| 2 | 0 | 2 | 28 | 0 | 30 | 93.3 |
| 3 | 0 | 0 | 0 | 14 | 14 | 100.0 |
| Total | 202 | 28 | 30 | 14 | 274 | 94.5 |

**Supplementary Table 10. Confusion matrix of combined score prediction on the prospective test dataset.**

| True label | Model prediction | | | | Total | Accuracy (%) |
|---|---|---|---|---|---|---|
| | 0 | 1 | 2 | 3 | | |
| 0 | 142 | 9 | 1 | 0 | 152 | 93.4 |
| 1 | 12 | 32 | 8 | 0 | 52 | 61.5 |
| 2 | 2 | 2 | 44 | 4 | 52 | 84.6 |
| 3 | 0 | 0 | 0 | 18 | 18 | 100.0 |
| Total | 152 | 52 | 52 | 18 | 274 | 86.1 |

**Supplementary Table 11. Performance of three SH score binary classification tasks on the external test dataset.** Data in parentheses are 95% confidence intervals.

| Task | AUC | PPV | NPV | Sensitivity (%) | Specificity (%) |
|---|---|---|---|---|---|
| 0 vs 1,2,3 | 0.940(0.920-0.960) | 0.815(0.784-0.846) | 0.909(0.882-0.936) | 87.1(83.0-91.2) | 86.4(83.7-89.0) |
| 0,1 vs 2,3 | 0.985(0.983-0.988) | 0.817(0.803-0.830) | 0.986(0.983-0.989) | 95.1(93.9-96.3) | 94.1(93.6-94.7) |
| 0,1,2 vs 3 | 0.979(0.974-0.984) | 0.613(0.577-0.649) | 0.996(0.994-0.997) | 95.6(93.8-97.4) | 94.3(93.4-95.1) |

**Supplementary Table 12. Performance of three vascularity score binary classification tasks on the external test dataset.** Data in parentheses are 95% confidence intervals.

| Task | AUC | PPV | NPV | Sensitivity (%) | Specificity (%) |
|---|---|---|---|---|---|
| 0 vs 1,2,3 | 0.998(0.995-1.000) | 0.954(0.939-0.969) | 0.994(0.992-0.996) | 98.5(98.0-99.0) | 98.2(97.6-98.8) |
| 0,1 vs 2,3 | 0.996(0.994-0.998) | 0.884(0.856-0.912) | 0.995(0.991-0.998) | 97.9(96.5-99.2) | 96.9(96.1-97.7) |
| 0,1,2 vs 3 | 0.988(0.974-1.000) | 0.796(0.734-0.858) | 0.998(0.996-1.000) | 98.1(95.7-100.6) | 97.3(96.2-98.3) |

**Supplementary Table 13. Accuracy and linearly weighted $\kappa$ comparison on the external test dataset.** Data in parentheses are 95% confidence intervals. $P$ values indicate comparison between RATING model and other methods.

| Metric | SH score | Vascularity score | Combined score | $P$ value |
|---|---|---|---|---|
| Accuracy(%) | | | | |
| RATING model | 82.9(78.5,87.0) | 96.2(93.9,98.3) | 85.0(80.5,89.1) | - |
| SH prediction using GSUS | 77.1(72.4,82.3) | 96.2(93.9,98.3) | 79.2(74.4,83.6) | $< 0.001$ |
| SH prediction using Doppler US | 80.5(76.1,85.3) | 96.2(93.9,98.3) | 82.6(78.2,87.0) | $< 0.001$ |
| Vascularity prediction using both modes | 82.9(78.5,87.0) | 91.5(88.1,94.5) | 83.3(79.2,87.4) | $< 0.001$ |
| w/o MULTITUDE | 74.4(69.2-79.6) | 94.7(93.2-96.2) | 76.1(70.8-81.5) | $< 0.001$ |
| Multi-classification | 70.0(65.4-74.6) | 89.6(88.8-90.5) | 71.9(67.6-76.1) | $< 0.001$ |
| Cascade [1] | - | - | 74.4(73.1-75.7) | $< 0.001$ |
| Linearly weighted $\kappa$ | | | | |
| RATING model | 0.832(0.789-0.919) | 0.957(0.932-0.953) | 0.857(0.817-0.897) | - |
| SH prediction using GSUS | 0.718(0.655-0.868) | 0.957(0.932-0.953) | 0.753(0.693-0.812) | $< 0.001$ |
| SH prediction using Doppler US | 0.796(0.747-0.803) | 0.957(0.932-0.953) | 0.829(0.784-0.874) | $< 0.001$ |
| Vascularity prediction using both modes | 0.832(0.789-0.875) | 0.908(0.877-0.939) | 0.844(0.804-0.884) | $< 0.001$ |
| w/o MULTITUDE | 0.741(0.690-0.793) | 0.940(0.923-0.956) | 0.771(0.721-0.821) | $< 0.001$ |
| Multi-classification | 0.680(0.633-0.727) | 0.863(0.842-0.885) | 0.721(0.679-0.763) | $< 0.001$ |
| Cascade [1] | - | - | 0.737(0.721-0.754) | $< 0.001$ |

**Supplementary Table 14. Confusion matrix of SH score prediction on the external test dataset.**

| True label | Model prediction | | | | Total | Accuracy (%) |
|---|---|---|---|---|---|---|
| | 0 | 1 | 2 | 3 | | |
| 0 | 163 | 10 | 1 | 0 | 174 | 93.7 |
| 1 | 11 | 32 | 13 | 0 | 56 | 57.1 |
| 2 | 1 | 0 | 24 | 13 | 38 | 63.2 |
| 3 | 0 | 0 | 2 | 23 | 25 | 92.0 |
| Total | 174 | 56 | 38 | 25 | 293 | 82.6 |

**Supplementary Table 15. Confusion matrix of vascularity score prediction on the external test dataset.**

| True label | Model prediction | | | | Total | Accuracy (%) |
|---|---|---|---|---|---|---|
| | 0 | 1 | 2 | 3 | | |
| 0 | 214 | 0 | 0 | 0 | 214 | 100.0 |
| 1 | 2 | 18 | 3 | 0 | 23 | 78.3 |
| 2 | 0 | 1 | 25 | 3 | 29 | 86.2 |
| 3 | 0 | 0 | 0 | 27 | 27 | 100.0 |
| Total | 214 | 23 | 29 | 27 | 293 | 96.9 |

**Supplementary Table 16. Confusion matrix of EOSS combined score prediction on the external test dataset.**

| True label | Model prediction | | | | Total | Accuracy (%) |
|---|---|---|---|---|---|---|
| | 0 | 1 | 2 | 3 | | |
| 0 | 163 | 9 | 1 | 0 | 173 | 94.2 |
| 1 | 11 | 32 | 11 | 0 | 54 | 59.3 |
| 2 | 0 | 1 | 22 | 11 | 34 | 64.7 |
| 3 | 0 | 0 | 1 | 31 | 32 | 96.9 |
| Total | 173 | 54 | 34 | 32 | 293 | 84.6 |

**Supplementary Table 17. Experience of radiologists in reader study and AI-assisted reader study.**

| Radiologist | US experience (year) | MKS-US experience (year) | EOSS training (time) |
|---|---|---|---|
| R1 | 8 | 3 | 7 |
| R2 | 5 | 2 | 5 |
| R3 | 4 | 1 | 3 |
| R4 | 15 | 3 | 5 |
| R5 | 13 | 3 | 5 |
| R6 | 13 | 3 | 5 |
| R7 | 4 | 3 | 7 |
| R8 | 5 | 1 | 3 |
| R9 | 13 | 3 | 5 |
| R10 | 5 | 1 | 3 |

**Supplementary Table 18. Accuracy of radiologists (R1-R10) and the average reader in reader study and AI-assisted reader study.** Data in parentheses are 95% confidence intervals. $P$ values indicate comparison between sole and AI-assisted combined score accuracy of each radiologist.

| Radiologist | | SH score (%) | Vascularity score (%) | Combined score (%) | $P$ value |
|---|---|---|---|---|---|
| R1 | sole | 40.1(34.7,46.0) | 82.5(78.1,87.2) | 39.8(34.3,45.6) | - |
| | AI-assisted | 71.2(65.7,76.6) | 83.9(79.9,88.3) | 70.8(65.7,76.3) | $< 0.001$ |
| R2 | sole | 24.1(19.0,29.2) | 76.3(71.2,81.4) | 27.0(21.9,32.5) | - |
| | AI-assisted | 49.3(43.1,55.1) | 81.4(77.0,85.8) | 48.5(42.3,54.4) | $< 0.001$ |
| R3 | sole | 36.9(31.4,42.3) | 80.7(75.9,85.0) | 39.8(33.9,45.6) | - |
| | AI-assisted | 77.0(71.9,82.1) | 90.9(87.6,94.2) | 81.8(77.0,86.5) | $< 0.001$ |
| R4 | sole | 37.6(32.1,43.4) | 81.0(75.9,85.4) | 40.5(34.3,46.4) | - |
| | AI-assisted | 48.5(43.1,54.4) | 87.6(83.2,91.2) | 52.9(47.8,58.8) | $< 0.001$ |
| R5 | sole | 49.3(43.8,55.5) | 79.9(75.2,84.7) | 50.7(45.3,56.9) | - |
| | AI-assisted | 68.6(63.5,74.1) | 86.1(81.8,90.5) | 71.5(66.8,77.0) | $< 0.001$ |
| R6 | sole | 31.8(26.3,37.6) | 81.8(77.0,86.5) | 36.1(30.7,42.0) | - |
| | AI-assisted | 73.0(67.9,77.7) | 87.6(83.6,91.6) | 76.3(70.8,81.4) | $< 0.001$ |
| R7 | sole | 48.9(43.1,54.7) | 78.1(73.4,83.2) | 48.2(42.7,54.0) | - |
| | AI-assisted | 59.1(53.6,65.0) | 82.8(78.1,87.2) | 60.2(54.7,66.1) | $< 0.001$ |
| R8 | sole | 29.6(24.5,34.7) | 81.0(76.3,85.4) | 33.6(28.1,39.4) | - |
| | AI-assisted | 33.9(28.5,39.4) | 80.7(75.5,85.8) | 35.8(30.3,41.2) | $< 0.001$ |
| R9 | sole | 39.8(34.3,45.6) | 79.9(75.2,84.3) | 39.8(34.7,46.0) | - |
| | AI-assisted | 68.2(62.8,73.7) | 87.6(83.9,91.2) | 70.4(65.0,75.9) | $< 0.001$ |
| R10 | sole | 58.0(52.2,63.9) | 82.5(77.7,86.9) | 58.0(52.6,63.9) | - |
| | AI-assisted | 73.0(67.5,78.5) | 85.8(81.8,89.8) | 71.9(66.4,77.0) | $< 0.001$ |
| Avg | sole | 39.6(34.1,45.3) | 80.4(75.6,85.0) | 41.4(35.8,47.2) | - |
| | AI-assisted | 62.2(56.8,67.7) | 85.4(81.2,89.6) | 64.0(58.7,69.5) | $< 0.001$ |

**Supplementary Table 19. Youden index of radiologists (R1-R10) and the average reader in reader study and AI-assisted reader study.** Data in parentheses are 95% confidence intervals. $P^{\#}$ values indicate comparison between sole and AI-assisted Youden index of each radiologist for predicting combined score 0 versus 1,2 and 3. $P^{*}$ values indicate comparison between sole and AI-assisted Youden index of each radiologist for predicting combined score 0,1 versus 2,3. $P^{\$}$ values indicate comparison between sole and AI-assisted Youden index of each radiologist for predicting combined score 0,1 and 2 versus 3.

| Radiologist | | 0 vs 1,2,3 | $P$ value$^{\#}$ | 0,1 vs 2,3 | $P$ value$^{*}$ | 0,1,2 vs 3 | $P$ value$^{\$}$ |
|---|---|---|---|---|---|---|---|
| R1 | sole | 0.189(0.122,0.270) | - | 0.637(0.533,0.741) | - | 0.438(0.192,0.666) | - |
| | AI-assisted | 0.597(0.506,0.691) | $< 0.001$ | 0.761(0.664,0.849) | $< 0.001$ | 0.548(0.309,0.786) | $< 0.001$ |
| R2 | sole | 0.102(0.048,0.161) | - | 0.322(0.217,0.422) | - | 0.351(0.106,0.596) | - |
| | AI-assisted | 0.377(0.291,0.460) | $< 0.001$ | 0.619(0.503,0.729) | $< 0.001$ | 0.592(0.350,0.825) | $< 0.001$ |
| R3 | sole | 0.137(0.056,0.218) | - | 0.615(0.504,0.732) | - | 0.421(0.176,0.655) | - |
| | AI-assisted | 0.785(0.704,0.856) | $< 0.001$ | 0.817(0.732,0.900) | $< 0.001$ | 0.731(0.520,0.910) | $< 0.001$ |
| R4 | sole | 0.160(0.096,0.229) | - | 0.576(0.488,0.670) | - | 0.620(0.393,0.836) | - |
| | AI-assisted | 0.314(0.233,0.399) | $< 0.001$ | 0.679(0.583,0.774) | $< 0.001$ | 0.735(0.524,0.913) | $< 0.001$ |
| R5 | sole | 0.324(0.240,0.412) | - | 0.565(0.448,0.673) | - | 0.373(0.148,0.612) | - |
| | AI-assisted | 0.576(0.484,0.674) | $< 0.001$ | 0.737(0.629,0.830) | $< 0.001$ | 0.758(0.545,0.940) | $< 0.001$ |
| R6 | sole | 0.291(0.211,0.389) | - | 0.443(0.360,0.534) | - | 0.572(0.369,0.742) | - |
| | AI-assisted | 0.698(0.605,0.779) | $< 0.001$ | 0.760(0.660,0.844) | $< 0.001$ | 0.786(0.598,0.946) | $< 0.001$ |
| R7 | sole | 0.301(0.211,0.388) | - | 0.486(0.365,0.611) | - | 0.437(0.192,0.659) | - |
| | AI-assisted | 0.423(0.343,0.512) | $< 0.001$ | 0.726(0.625,0.826) | $< 0.001$ | 0.699(0.477,0.908) | $< 0.001$ |
| R8 | sole | 0.082(0.033,0.137) | - | 0.489(0.377,0.596) | - | 0.648(0.427,0.860) | - |
| | AI-assisted | 0.166(0.104,0.236) | $< 0.001$ | 0.489(0.391,0.594) | 0.659 | 0.410(0.172,0.641) | $< 0.001$ |
| R9 | sole | 0.183(0.107,0.261) | - | 0.479(0.363,0.599) | - | 0.671(0.442,0.883) | - |
| | AI-assisted | 0.583(0.479,0.685) | $< 0.001$ | 0.616(0.499,0.726) | $< 0.001$ | 0.802(0.616,0.969) | $< 0.001$ |
| R10 | sole | 0.492(0.400,0.594) | - | 0.592(0.473,0.719) | - | 0.386(0.152,0.629) | - |
| | AI-assisted | 0.678(0.590,0.771) | $< 0.001$ | 0.480(0.360,0.601) | $< 0.001$ | 0.536(0.294,0.769) | $< 0.001$ |
| Avg | sole | 0.226(0.153,0.306) | - | 0.520(0.413,0.630) | - | 0.492(0.260,0.714) | - |
| | AI-assisted | 0.520(0.434,0.606) | $< 0.001$ | 0.668(0.565,0.767) | $< 0.001$ | 0.660(0.440,0.861) | $< 0.001$ |

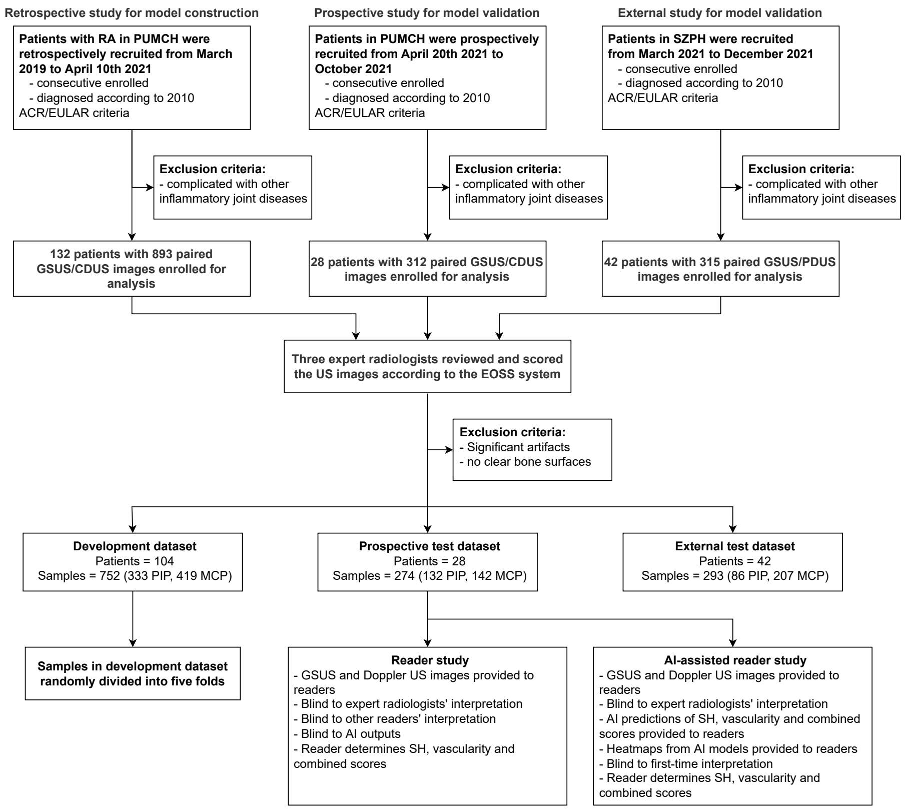

**Supplementary Figure 1. Overview of the retrospective, prospective and external study workflow.**

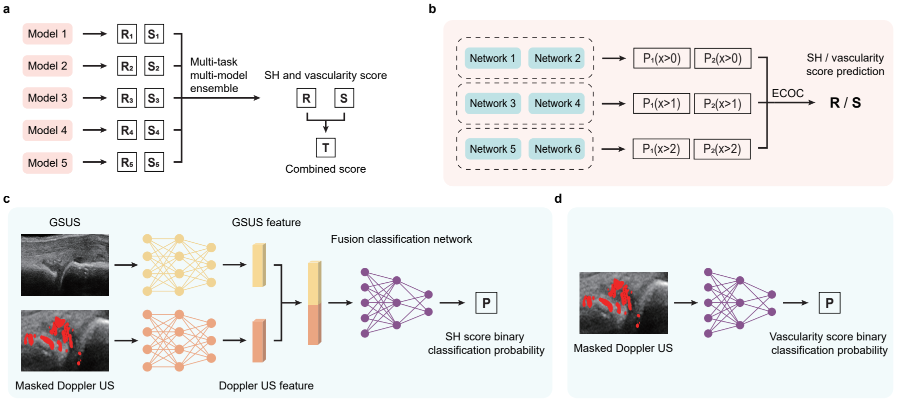

**Supplementary Figure 2. Structure of the RATING model. a**, Five models separately predict the SH score and the vascularity score, and all the predictions are comprehensively analyzed using multi-task multi-model ensemble (MULTITUDE) to yield final predictions. **b**, Both SH scoring module and vascularity scoring module are composed of six networks for three binary classification tasks, and SH score or vascularity score prediction is obtained from six binary predictions using ECOC. **c**, Each GS-Doppler feature fusion network extracts feature vectors from GSUS and Doppler US images, fuses them and predicts SH score binary classification probabilities. **d**, Each vascularity score binary classification network predicts vascularity score binary classification probabilities using Doppler US images.

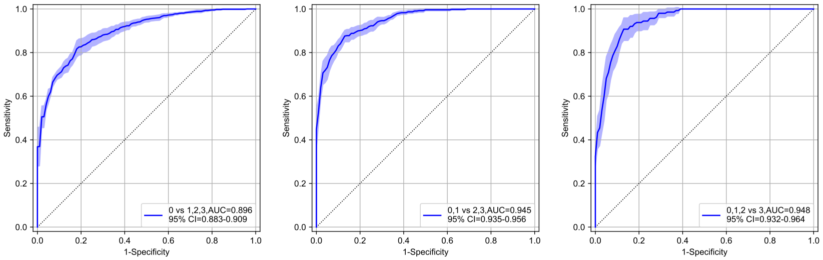

**(a)** 0 vs 1,2,3 **(b)** 0,1 vs 2,3 **(c)** 0,1,2 vs 3

**Supplementary Figure 3. Performance of three SH score binary classification tasks on the development dataset.** ROC curves of SH score classification between 0 and 1,2,3 (**a**), between 0,1 and 2,3 (**b**), and between 0,1,2 and 3 (**c**). The shaded regions represent the 95% confidence intervals. Data in parentheses are 95% confidence intervals.

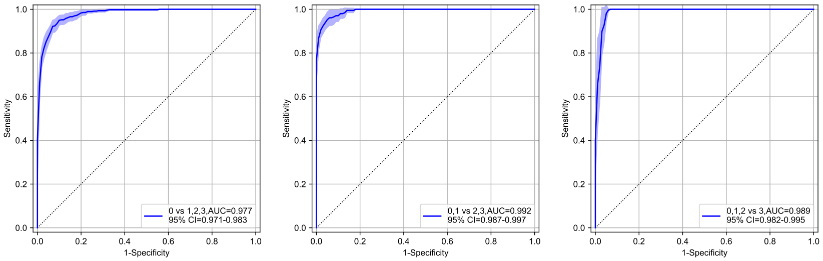

**(a)** 0 vs 1,2,3 **(b)** 0,1 vs 2,3 **(c)** 0,1,2 vs 3

**Supplementary Figure 4. Performance of three vascularity score binary classification tasks on the development dataset.** ROC curves of SH score classification between 0 and 1,2,3 (**a**), between 0,1 and 2,3 (**b**), and between 0,1,2 and 3 (**c**). The shaded regions represent the 95% confidence intervals. Data in parentheses are 95% confidence intervals.

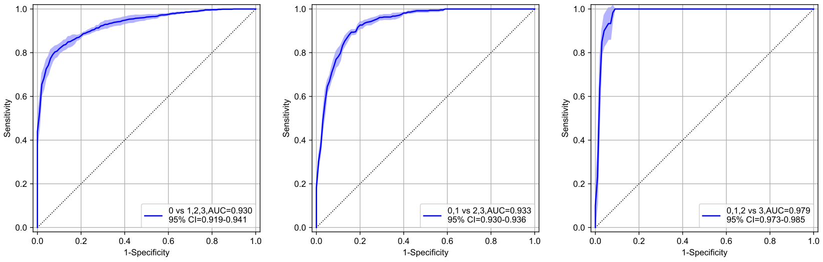

**(a)** 0 vs 1,2,3 **(b)** 0,1 vs 2,3 **(c)** 0,1,2 vs 3

**Supplementary Figure 5. Performance of three SH score binary classification tasks on the prospective test dataset.** ROC curves of SH score classification between 0 and 1,2,3 (**a**), between 0,1 and 2,3 (**b**), and between 0,1,2 and 3 (**c**). The shaded regions represent the 95% confidence intervals. Data in parentheses are 95% confidence intervals.

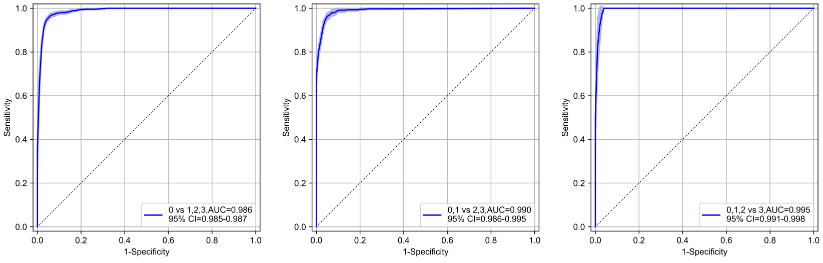

**(a)** 0 vs 1,2,3 **(b)** 0,1 vs 2,3 **(c)** 0,1,2 vs 3

**Supplementary Figure 6. Performance of three vascularity score binary classification tasks on the prospective test dataset.** ROC curves of SH score classification between 0 and 1,2,3 (**a**), between 0,1 and 2,3 (**b**), and between 0,1,2 and 3 (**c**). The shaded regions represent the 95% confidence intervals. Data in parentheses are 95% confidence intervals.

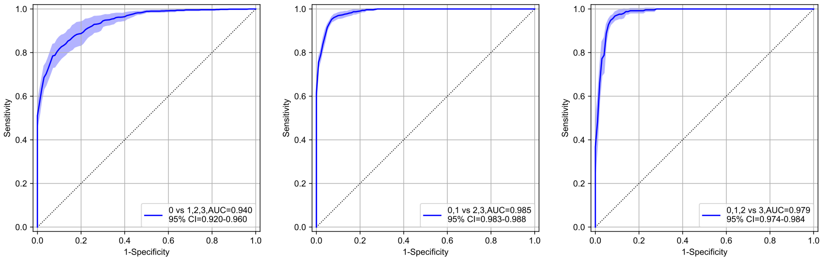

**(a)** 0 vs 1,2,3 **(b)** 0,1 vs 2,3 **(c)** 0,1,2 vs 3

**Supplementary Figure 7. Performance of three SH score binary classification tasks on the external test dataset.** ROC curves of vascularity score classification between 0 and 1,2,3 (**a**), between 0,1 and 2,3 (**b**), and between 0,1,2 and 3 (**c**). The shaded regions represent the 95% confidence intervals. Data in parentheses are 95% confidence intervals.

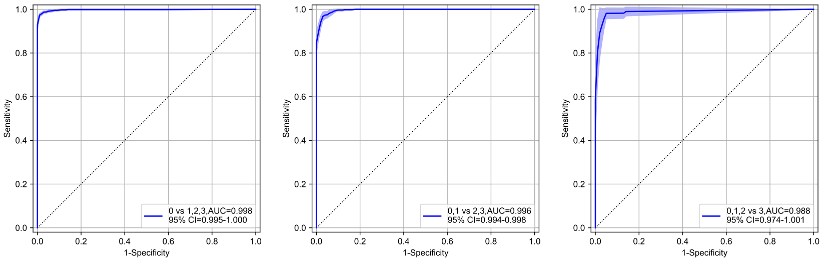

**(a)** 0 vs 1,2,3 **(b)** 0,1 vs 2,3 **(c)** 0,1,2 vs 3

**Supplementary Figure 8. Performance of three vascularity score binary classification tasks on the external test dataset.** ROC curves of vascularity score classification between 0 and 1,2,3 (**a**), between 0,1 and 2,3 (**b**), and between 0,1,2 and 3 (**c**). The shaded regions represent the 95% confidence intervals. Data in parentheses are 95% confidence intervals.

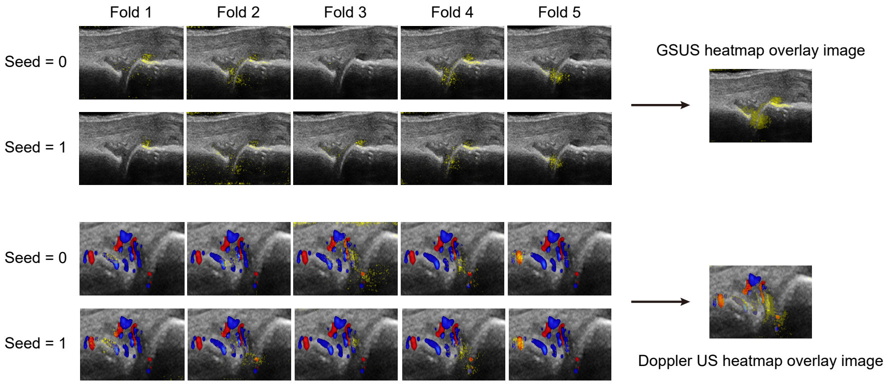

**Supplementary Figure 9. Illustration of heatmap overlay image generation process.** For each GSUS and Doppler US image, heatmaps are generated from ten GS-Doppler feature fusion networks and averaged. The heatmaps are colorized by yellow and overlaid on the original US images.
